# Creating an 11-year longitudinal substance use harm cohort from linked health and census data to analyze social drivers of health

**DOI:** 10.1101/2024.02.14.24302828

**Authors:** Anousheh Marouzi, Charles Plante, Barbara Fornssler

## Abstract

Research on substance use harm in Saskatchewan has faced challenges due to an absence of linked data to analyze and report on the social drivers of substance use harm. This study uses the Canadian Census Health and Environment Cohorts (CanCHECs) 2006 to create, describe, and validate a cohort of Saskatchewan residents focusing on substance use. We achieved validation by comparing our descriptive findings with those from other Canadian studies on substance use. The second objective of this study was to underscore the potential CanCHEC holds in studying substance use, especially by bridging the gap in data concerning the linkage of social determinants of health and administrative health data. Additionally, to facilitate further research using this rich national data source, we share our Stata do-file, providing a detailed walkthrough for creating national or provincial substance use cohorts.

**About the Research Department:** The Saskatchewan Health Authority Research Department leads collaborative research to enhance Saskatchewan’s health and healthcare. We provide diverse research services to SHA staff, clinicians, and team members, including surveys, study design, database development, statistical analysis, and assistance with research funding. We also spearhead our own research programs to strengthen research and analytic capability and learning within Saskatchewan’s health system.

**Disclaimer:** This working paper is for discussion and comment purposes. It has not been peer-reviewed nor been subject to review by Research Department staff or executives. Any opinions expressed in this paper are those of the author(s) and not those of the Saskatchewan Health Authority.

**Suggested Citation:** Marouzi Anousheh, Plante Charles, and Fornssler Barbara. 2024. “Creating an 11-year longitudinal substance use harm cohort from linked health and census data to analyze social drivers of health.” MedRxiv.

**Extended Abstract:** *Background:* Research on substance use harm in Saskatchewan has been hampered by an absence of linked data to analyze and report on the social drivers of substance use harm. This study aims to create, describe, and validate a cohort of Saskatchewan residents by linking their sociodemographic data to their health outcomes using line-level data made available by Statistics Canada’s Research Data Centres (RDC) program.

*Methods:* We used Canadian Census Health and Environment Cohorts (CanCHECs) 2006 to create a cohort of Saskatchewanians followed from 2006 to 2016. We linked sociodemographic information of the 2006 Census (long-form) respondents to their hospitalization data captured in the Discharge Abstract Database (DAD) (2006 to 2016) and their mortality records in the Canadian Vital Statistics Death Database (CVSD) (2006 to 2016.) We developed an algorithm to identify Saskatchewanians who experienced a substance use harm event. We validated the cohort by comparing our descriptive findings with those from other Canadian studies on substance use.

*Results:* We used CanCHEC, a national data resource, whereas most previous studies have used provincial data resources. Despite this difference in constructing the cohorts, our results showed trends consistent with previous studies, including an overrepresentation of individuals with lower socioeconomic status within the PESUH group. Similar to other Canadian studies, our results indicate an increasing rate of substance use harm from 2006 to 2016. To facilitate further research using CanCHEC, we share our Stata do-file, providing a detailed walkthrough so other researchers can create national or provincial substance use cohorts.

*Conclusion:* Using CanCHEC to create substance use cohorts will enable health researchers to provide a province-wide, population-level, and longitudinal perspective on substance use harm. This comprehensive view is crucial in effectively contextualizing smaller-scale and local studies, allowing us to disentangle the “fundamental causes” of health within the region.

*Key Messages:*

- CanCHEC provides researchers with an excellent opportunity to measure and examine health inequalities across socioeconomic and ethnocultural dimensions for different periods and locations in Canada.
- There has been a steady increase in people who experienced substance use harm in Saskatchewan, from 2006 to 2016.
- People who experienced substance use harm between 2006 and 2016 were overrepresented among individuals with an education level below high school, those in the lowest income quintile, residents of rural areas, and Indigenous population.
- This study provides a Stata do-file, including a detailed walkthrough for using CanCHEC to create national or provincial substance use cohorts.

## Introduction

Research on social drivers of substance use harm (SUH) in Saskatchewan has been hampered by an absence of infrastructure that supports routine access to and analysis of linked health administrative and sociodemographic data. This limitation has prevented us from producing the kind of province-wide population level and overtime portrait of substance use in Saskatchewan that can be used to effectively situate smaller scale and local studies, untangle the “fundamental causes”^1,2^ of health, inform decision making, and improve service delivery and population health in the province.^3,4^

Understanding social determinants of substance use harm is vital, given the profound influence they have on an individual’s trajectory with using substances. Substance use is not simply a lifestyle “choice.” Past experiences, such as families, neighbourhoods, income, education, employment, and occupation, can play significant parts in future substance use-related harm.^5^ Moreover, these same social determinants also mediate access to care.^6,7^ Furthermore, since socioeconomic status is often spatially concentrated, there is a geographical variation in substance use harm rates across Canada.^5^ Recent Canadian research has found that substance use harm is more concentrated and visible in urban areas, rates are actually higher in rural areas.^8^ Additionally, mental health and substance use are known drivers of high-cost healthcare use in Canada.^9–11^ To curb related healthcare spending, it is crucial to identify factors amenable to public health actions that potentially can reduce these costs.^9^ This also underscores the need for a comprehensive data infrastructure enabling researchers to examine related factors, such as socioeconomic status.

There have been efforts to create provincial cohorts to delve into substance use harm in regions like British Columbia^12^ and Saskatchewan.^13^ However, these studies had limited ability to provide socio-economic insights due to the absence of linked health data to social information. However, this gap in the information can be addressed by using the rich national databases, such as the Canadian Census Health and Environment Cohorts (CanCHECs), accessible at Research Data Centres (RDC)^14^ throughout the country. The RDC facilitates research that uses sensitive microdata within a secure research environment managed by Statistics Canada.^14^

In response to the increased demand for including social data in epidemiology research, Statistics Canada has launched the CanCHECs project to relate social information gathered through its bi-decennial censuses (long form) and National Household Survey (NHS) to health administrative and registry databases at the individual level.^15^ This includes the Discharge Abstract Database (DAD), Canadian Vital Statistics Death Database (CVSD), National Ambulatory Care Reporting System (NACRS), and Canadian Cancer Registry (CCR). CanCHEC also links these data to Canada Revenue Agency (CRA) tax files to trace where people have historically resided, making the data ideal for environmental exposure research.^15^ As of August 2023, CanCHEC includes six cycles spanning from 1991 to 2016.^16^

CanCHEC provides researchers with an excellent opportunity to measure and examine health inequalities across socioeconomic and ethnocultural dimensions for different periods and locations in Canada.^15^ Yet, to date, only one study has used CanCHEC to examine the social drivers of substance use harm in Canada.^17^ Carrière et al. used it to describe the socioeconomic characteristics of those experiencing hospitalizations due to opioid poisoning between 2011 and 2016.^17^

The main objective of this study is to create, describe, and validate a substance use cohort of Saskatchewanians, developed using CanCHEC. To validate this cohort, we compared our descriptive findings against those from previous studies on substance use in Canada, such as the British Columbia^12^ and Saskatchewan^13^ substance use cohorts. The second objective was to highlight the potential CanCHEC holds for a micro-level linkage of socioeconomic and ethnocultural data to health administrative data. Figure 1 details how we created our provincial cohort, which includes a key subgroup of people who experienced substance use harm (PESUH), from CanCHEC datasets in three steps. We created our provincial cohort in Step Two by extracting Saskatchewan residents from a national cohort that was created in Step One. Cohorts for other provinces could readily be created by extracting their residents instead. Or, a national cohort could be created by skipping Step Two.

**Figure 1.**
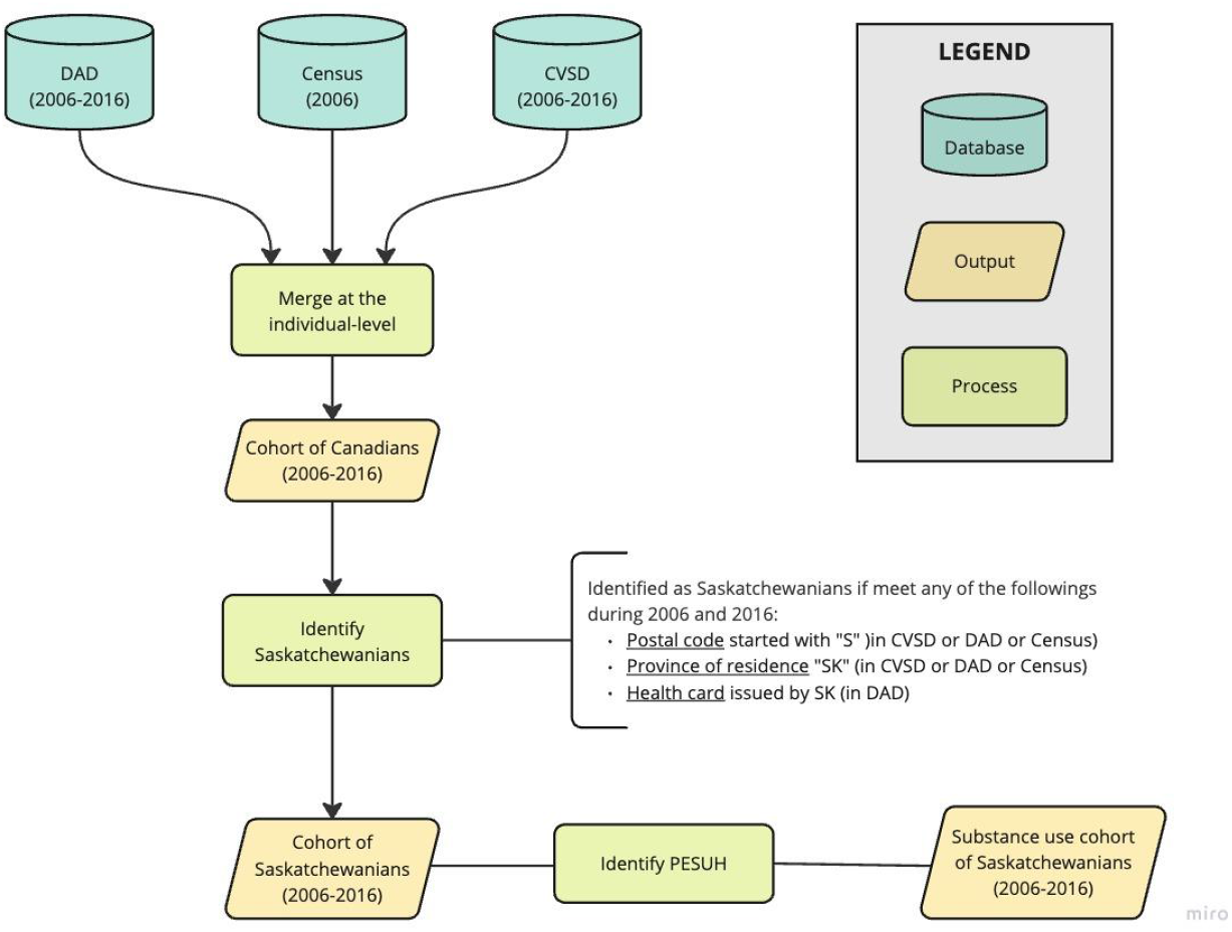
Creating the Substance Use Cohort of Saskatchewanians. Notes: DAD: Discharge Abstract Dataset; CVSD: Canadian Vital Statistics Death Database; PESUH: People who have experienced substance use harm.

To facilitate working with the complicated data structure of CanCHEC, we provide our Stata do-file, which includes a step-by-step guide for creating national/provincial substance use cohorts. The cohort produced by this do-file offers a great opportunity to researchers, knowledge users, and patient-led research initiatives working to better understand substance use harm at the national, provincial, and smaller scale and local levels.

## Methods

### Data sources

We used CanCHEC 2006 to create a cohort of Saskatchewanians and explore substance use harm among them. “The CanCHECs are a series of population-based, probabilistically linked datasets that combine data from respondents to the long-form census or 2011 National Household Survey (NHS) with administrative health data (e.g., mortality, cancer incidence, hospitalizations, emergency ambulatory care) and annual postal code history.”^15^ We chose the 2006 cycle because it gives the opportunity to prospectively follow the respondents over the longest number of years, 11 years, compared to 6 years in the 2011 cycle and 5 years in the 2016 cycle.^16^ CanCHEC also provides the opportunity to retrospectively follow the census respondents. For example, the 2016 cycle enables a 17-year retrospective follow-up.

However, retrospective follow-up leads to survivorship bias, a form of selection bias.^18,19^ In the 2016 CanCHEC, respondents have to have lived to complete the 2016 census for their retrospective trajectories to be included, and therefore people deceased in previous years had no chance of being selected in the cohort. The longer our retrospective follow-up and the more likely a population is to have not survived to the census, a particularly acute risk for individuals who have suffered substance use harms, the greater this survivorship bias is likely to be.

The CanCHEC 2006 includes usual residents of Canada as of the census date, capturing both permanent and non-permanent residents who responded to the 2006 long-form Census,^20^ but excluding institutionalized populations (e.g., those living in nursing homes, penitentiaries, and group homes.)^15^ Of the 6,463,927 long-form census respondents in 2006, 90.8% (5,871,337 records) qualified for CanCHEC.^15^ These records are linked to 436,407 Canadian Vital Statistics Death Data (CVSD)^21^ records (2006 to 2016) and 8,923,516 Discharge Abstract Database (DAD)^22^ records (2000/2001 to 2016/2017.)^15^ For this study, we linked Census respondents eligible for CanCHEC to 2006-2016 hospitalization and death data recorded in DAD and CVSD. We excluded the National Ambulatory Care Reporting System (NACRS)^23^ due to its inconsistent coverage in Saskatchewan between 2006 and 2016.

### Cohort creation

The substance use cohort of Saskatchewanians was created in three steps (Figure 1): (1) creating a national cohort by linking sociodemographic and administrative health data, (2) identifying Saskatchewanians in the national cohort and creating the provincial cohort, and (3) identifying people who experienced substance use-related hospitalization and deaths.

#### 1. Linking hospitalization, mortality, and sociodemographic data

CanCHEC datasets have key files enabling the linkage of Census data to administrative health data. Appendix A illustrates the relationship between CanCHEC 2006’s datasets used in this study to link the sociodemographic data available in the Census to hospitalization records in DAD (2006 to 2016) and mortality data in CVSD (2006 to 2016,) at the individual level. The final output at this step was a cohort of Canadians whose hospitalization and mortality information were tracked from 2006 to 2016.

#### 2. Identifying Saskatchewanians

We used the national cohort produced in the previous step to create a provincial cohort that includes people who resided in Saskatchewan at any point in time between 2006 and 2016. We defined a set of criteria to identify these people. We considered a person Saskatchewanian if the postal code for their place of residence started with “S” in CVSD or DAD or Census; or if their province of residence was Saskatchewan in CVSD or DAD or Census; or if their health card was issued by Saskatchewan as recorded in DAD. In other words, a person who lived in British Columbia in 2006 but moved to Saskatchewan in 2008 and thereafter experienced a substance use harm event recorded in the CVSD or DAD would appear in our cohort. For a small number of records, while their postal codes started with “S,” their province of residence was Alberta in the Census records. After further investigation and mapping the dissemination area corresponding to these postal codes in QGIS, we found out that these postal codes are related to Onion Lake, a reservation located on the boundary of Saskatchewan and Alberta. We decided to include the individuals who live in the Alberta section of Onion Lake in our cohort of Saskatchewanians. At the end of this step, individuals who were identified as Saskatchewanians were included in the cohort and the remaining records were excluded.

#### 3. Identifying people who experienced substance use harm

We used the International Statistical Classification of Diseases and Related Health program (ICD-10)^24^ codes to identify hospitalizations or deaths that happened due to SUH. The case-finding algorithm used to identify SUH events is provided in Appendix B. This algorithm was developed based on methodologies applied by other researchers.^12,25–28^ We did not include opioid-related adverse drug reactions (Y45.0) as we were not interested in the harms resulting from the adverse effects of prescribed medications. It should also be noted that this list only contains harms that are 100% attributable to substance use, and therefore harms partially attributable to substance use (e.g., cancer, stroke) are excluded from this analysis.

After creating the cohort of Saskatchewanians, we identified two subpopulations of people who experienced substance use harm (PESUH) and people who have not experienced substance use harm (NPESUH) within the cohort. People were categorized in the PESUH group if they experienced at least one SUH event (i.e., hospitalization or death) between 2006 and 2016. Anyone who was not identified as PESUH was assigned to the NPESUH group.

### Descriptive analysis

We described the cohort of Saskatchewanians and its subgroups, PESUH and NPESUH, by age, sex, income, region, race, occupation, employment status, education, and number of hospitalizations (See Appendix C for more detail.) We used the after-tax household income variable captured by the census and adjusted it to the household size. The adjusted after-tax household income was then used to create income quintiles at the provincial level. We also plotted the annual rates of PESUH per 100,000 people, from 2006 to 2016, to present a picture of substance use harm trends in Saskatchewan, broken down by substance categories. In calculating the annual rates, we took the number of PESUH in a year as the numerator, and the number of people in the cohort who were alive in that year as the denominator. We applied the CanCHEC weight variable in all the calculations to produce estimates representative of the non-institutional population in Saskatchewan at the time of Census 2006. Moreover, for privacy considerations, we rounded the numerators and denominators to the nearest multiple of five before calculating the rates and percentages, following the Statistics Canada Research Data Centre (RDC) vetting rules.

Data was accessed via the Saskatchewan Research Data Centre on the University of Saskatchewan campus (SKY-RDC).^14^ Ethics exemption was obtained from the Saskatchewan Health Authority Research Ethics Board (REB-22-20).

### Validation analysis

We validated our cohort by comparing its descriptive characteristics with those from other Canadian studies on substance use. In doing so, we profiled the PESUH group using various social determinants of health and compared our results with what has been found by other researchers using different data sources. This validation was crucial, as CanCHEC’s dataset structure is fundamentally different compared to those used in other studies. CanCHEC is a federal data repository linking various national datasets at the individual level. The secure research environment provided by RDCs has made it possible for the researchers to access such line-level data. However, Canadian researchers often use provincial health data repositories that are not linked to social information and are limited to only one province.

## Results

### Identification of people who experienced substance use harm

Out of the cohort population of 228,000, approximately 7,000 individuals (3.07%) experienced at least one SUH event from 2006 to 2016 (PESUH.) These individuals were further categorized based on the type of substances recorded as a cause of their hospitalization or death. Notably, across all substance categories, over 95% of these individuals were identified exclusively through their hospitalization records. This indicates that while they were hospitalized due to that substance during the follow-up period, there were no recorded deaths related to that substance. A very small portion of the PESUH in the cohort were identified through both hospitalization and death records and an even smaller fraction solely via vital statistics death data. The data sources used for identifying PESUH are presented in Figure 2.

**Figure 2.**
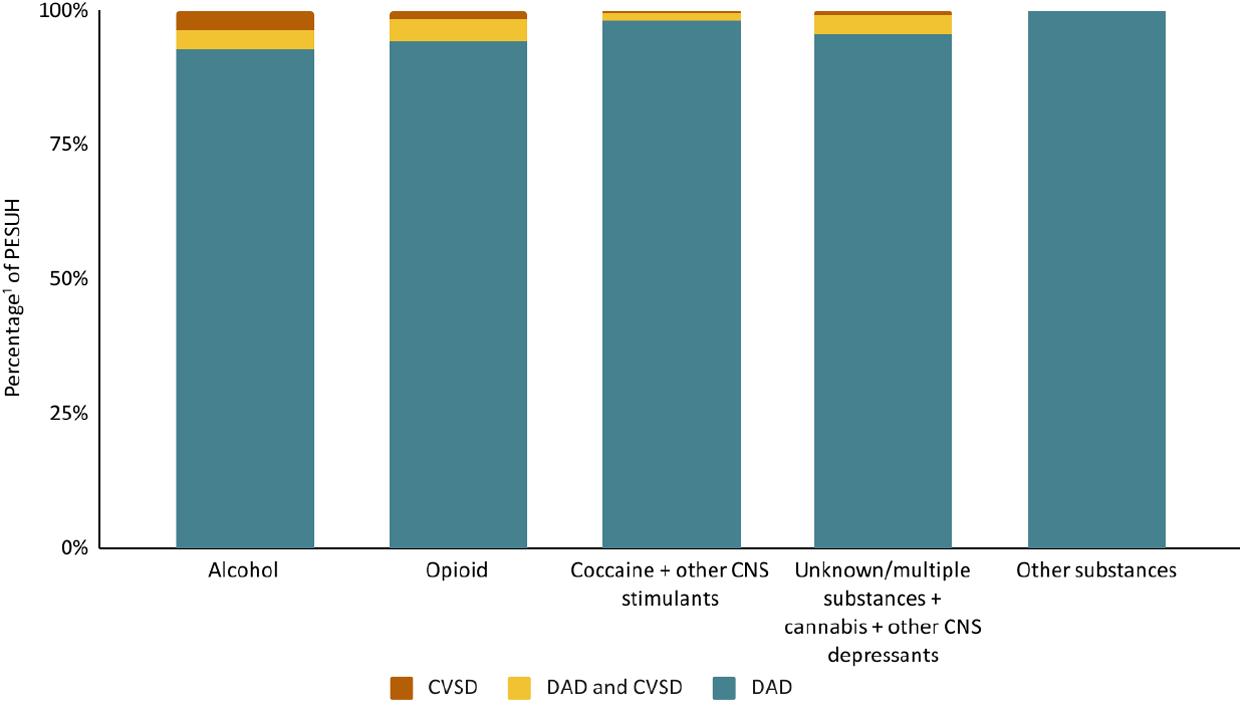
Identification of people who experienced substance use harm. Notes: We aggregated cocaine with other CVS stimulants and unknown/multiple substances with cannabis and other CNS depressants due to the low numbers of people falling in the CVSD categories. The RDC vetting rules do not allow releasing percentages with a numerator or denominator smaller than 5. PESUH: people who experienced substance use harm. CNS: central nervous system. ^1^Numerator: weighted number of PESUH identified by data source; denominator: weighted number of PESUH.

**Figure 3.**
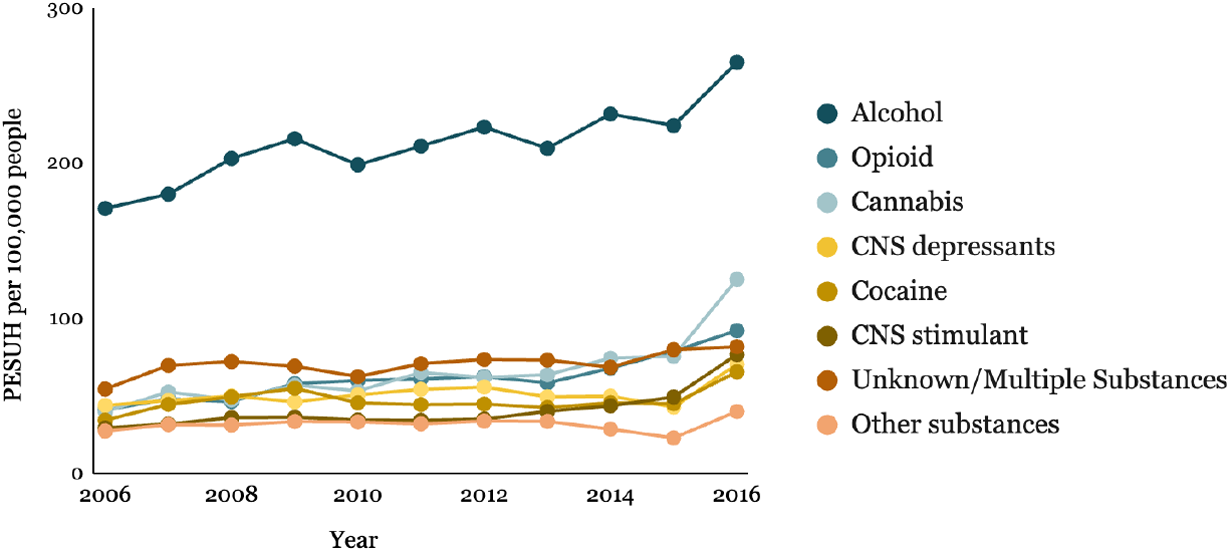
Trends of the number of people who have experienced substance use harm in Saskatchewan by substance category, 2006 to 2016. Notes: PESUH: people who experienced substance use harm.

The proportion of PESUH found through DAD and CVSD data is similar to what has been reported by Homayra et al. concerning the BC cohort.^12^ By comparing our cohort to the BC cohort,^12^ we can estimate that our PESUH group might have been two to three times larger had we accessed and incorporated the Medical Services Plan (MSP) records for identifying people who experienced substance use harm.

### Trends by SUH category, 2006 to 2016

Our findings indicate an increase in rates of PESUH per 100,000 population in Saskatchewan from 2006 to 2016. This uptrend spanned across all eight substance categories examined in this study. While there has been steady growth over these years, a huge jump in the PESUH rate was evident between 2015 and 2016. The data table on these trends is provided in Appendix D. We also calculated the age-standardized rates to ensure that these trends are not affected by the fact that our cohort is getting older through the 11 years of follow-up. The trends for age-standardized rates were similar to the non-age-standardized rates trends (not reported), meaning that the aging of the cohort was not a factor here.

We found that in Saskatchewan, the rates of PESUH for alcohol and cannabis were 265 and 125 per 100,000 population in 2016, compared to 224 and 75 per 100,000 population in 2015. According to CIHI’s report, the overall age-standardized rate of SUH-related hospitalization among youth aged 10 to 24 was 667 per 100,000 people in Saskatchewan in 2017-2018, which was much higher than the national level in that year (364 per 100,000 population.)^8^ Within Saskatchewan, the rates for cannabis and alcohol were 345 and 259, respectively. In contrast, the national rates for these substances were 141 and 95.^8^ A potential explanation for the spike in cannabis-related hospitalizations may be the increase in reporting cannabis use among youth after the legalization of it in Canada.^29^

### Number of hospitalizations

Our analysis indicates that people who have experienced at least one SUH event between 2006 and 2016 were far more likely to be admitted to the hospital during this timeframe. While 45.8% of the PESUH group had 2 to 5 hospital admissions, 40.0% were admitted 6 times or more. In contrast, the rest of the population exhibited lower hospitalization rates, with 31.8% admitted 2 to 5 times and only 11.5% admitted 6 times or more in the same period. However, it should be noted that since a primary criterion for inclusion in the PESUH group was hospitalization due to substance use, some of this gap may be due to this inclusion criterion.

### Socio-demographic characteristics

Table 1 provides the socio-demographic characteristics of the entire cohort and its two subgroups in 2006. Overall, modal individuals who experienced substance use harm events during the follow-up period were young (median age of 32), white^1^ (56.9%), working (38.7%) or seeking employment (9.3%), held at least a high school degree (38.7%), and resided in urban areas (63.4%) in 2006 (study baseline.) However, the PESUH group had an overrepresentation of individuals with less than a high school education (38.7% in PESUH vs. 23.7% in NPESUH), those in the low-income quintile (41.5% in PESUH vs. 19.5% in NPESUH), residents of rural areas (36.6% in PESUH vs. 34.2% in NPESUH), and individuals of Indigenous ethnicity (41.6% in PESUH vs. 13.8% in NPESUH.)

**Table 1:**
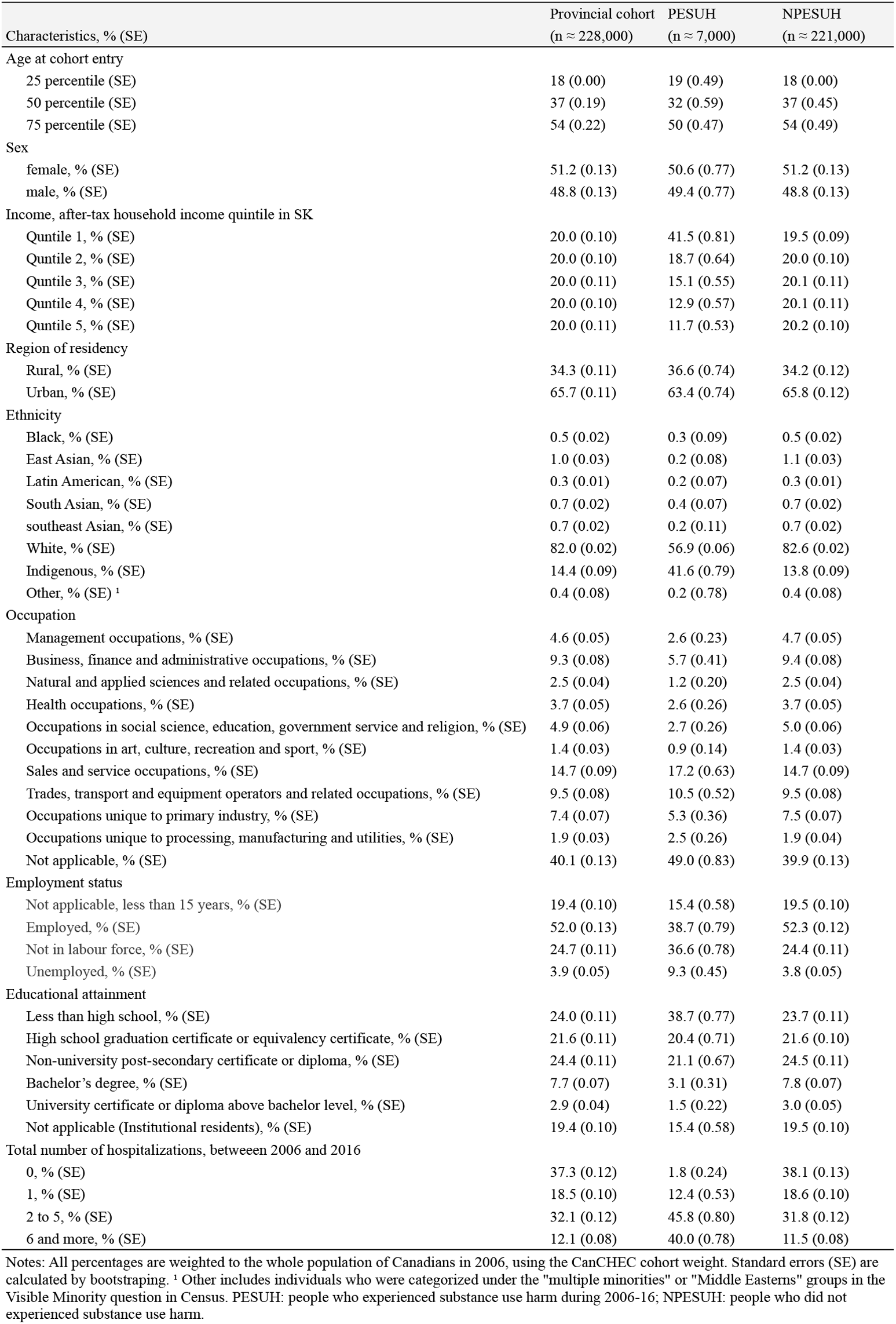
Sociodemographic characteristics of Census 2006 respondents, by having experience of substance use harm between 2006 to 2016.

| Characteristics, % (SE) | Provincial cohort<br>(n ≈ 228,000) | PESUH<br>(n ≈ 7,000) | NPESUH<br>(n ≈ 221,000) |
| --- | --- | --- | --- |
| Age at cohort entry |  |  |  |
| 25 percentile (SE) | 18 (0.00) | 19 (0.49) | 18 (0.00) |
| 50 percentile (SE) | 37 (0.19) | 32 (0.59) | 37 (0.45) |
| 75 percentile (SE) | 54 (0.22) | 50 (0.47) | 54 (0.49) |
| Sex |  |  |  |
| female, % (SE) | 51.2 (0.13) | 50.6 (0.77) | 51.2 (0.13) |
| male, % (SE) | 48.8 (0.13) | 49.4 (0.77) | 48.8 (0.13) |
| Income, after-tax household income quintile in SK |  |  |  |
| Quntile 1, % (SE) | 20.0 (0.10) | 41.5 (0.81) | 19.5 (0.09) |
| Quntile 2, % (SE) | 20.0 (0.10) | 18.7 (0.64) | 20.0 (0.10) |
| Quntile 3, % (SE) | 20.0 (0.11) | 15.1 (0.55) | 20.1 (0.11) |
| Quntile 4, % (SE) | 20.0 (0.10) | 12.9 (0.57) | 20.1 (0.11) |
| Quntile 5, % (SE) | 20.0 (0.11) | 11.7 (0.53) | 20.2 (0.10) |
| Region of residency |  |  |  |
| Rural, % (SE) | 34.3 (0.11) | 36.6 (0.74) | 34.2 (0.12) |
| Urban, % (SE) | 65.7 (0.11) | 63.4 (0.74) | 65.8 (0.12) |
| Ethnicity |  |  |  |
| Black, % (SE) | 0.5 (0.02) | 0.3 (0.09) | 0.5 (0.02) |
| East Asian, % (SE) | 1.0 (0.03) | 0.2 (0.08) | 1.1 (0.03) |
| Latin American, % (SE) | 0.3 (0.01) | 0.2 (0.07) | 0.3 (0.01) |
| South Asian, % (SE) | 0.7 (0.02) | 0.4 (0.07) | 0.7 (0.02) |
| southeast Asian, % (SE) | 0.7 (0.02) | 0.2 (0.11) | 0.7 (0.02) |
| White, % (SE) | 82.0 (0.02) | 56.9 (0.06) | 82.6 (0.02) |
| Indigenous, % (SE) | 14.4 (0.09) | 41.6 (0.79) | 13.8 (0.09) |
| Other, % (SE) <sup>1</sup> | 0.4 (0.08) | 0.2 (0.78) | 0.4 (0.08) |
| Occupation |  |  |  |
| Management occupations, % (SE) | 4.6 (0.05) | 2.6 (0.23) | 4.7 (0.05) |
| Business, finance and administrative occupations, % (SE) | 9.3 (0.08) | 5.7 (0.41) | 9.4 (0.08) |
| Natural and applied sciences and related occupations, % (SE) | 2.5 (0.04) | 1.2 (0.20) | 2.5 (0.04) |
| Health occupations, % (SE) | 3.7 (0.05) | 2.6 (0.26) | 3.7 (0.05) |
| Occupations in social science, education, government service and religion, % (SE) | 4.9 (0.06) | 2.7 (0.26) | 5.0 (0.06) |
| Occupations in art, culture, recreation and sport, % (SE) | 1.4 (0.03) | 0.9 (0.14) | 1.4 (0.03) |
| Sales and service occupations, % (SE) | 14.7 (0.09) | 17.2 (0.63) | 14.7 (0.09) |
| Trades, transport and equipment operators and related occupations, % (SE) | 9.5 (0.08) | 10.5 (0.52) | 9.5 (0.08) |
| Occupations unique to primary industry, % (SE) | 7.4 (0.07) | 5.3 (0.36) | 7.5 (0.07) |
| Occupations unique to processing, manufacturing and utilities, % (SE) | 1.9 (0.03) | 2.5 (0.26) | 1.9 (0.04) |
| Not applicable, % (SE) | 40.1 (0.13) | 49.0 (0.83) | 39.9 (0.13) |
| Employment status |  |  |  |
| Not applicable, less than 15 years, % (SE) | 19.4 (0.10) | 15.4 (0.58) | 19.5 (0.10) |
| Employed, % (SE) | 52.0 (0.13) | 38.7 (0.79) | 52.3 (0.12) |
| Not in labour force, % (SE) | 24.7 (0.11) | 36.6 (0.78) | 24.4 (0.11) |
| Unemployed, % (SE) | 3.9 (0.05) | 9.3 (0.45) | 3.8 (0.05) |
| Educational attainment |  |  |  |
| Less than high school, % (SE) | 24.0 (0.11) | 38.7 (0.77) | 23.7 (0.11) |
| High school graduation certificate or equivalency certificate, % (SE) | 21.6 (0.11) | 20.4 (0.71) | 21.6 (0.10) |
| Non-university post-secondary certificate or diploma, % (SE) | 24.4 (0.11) | 21.1 (0.67) | 24.5 (0.11) |
| Bachelor's degree, % (SE) | 7.7 (0.07) | 3.1 (0.31) | 7.8 (0.07) |
| University certificate or diploma above bachelor level, % (SE) | 2.9 (0.04) | 1.5 (0.22) | 3.0 (0.05) |
| Not applicable (Institutional residents), % (SE) | 19.4 (0.10) | 15.4 (0.58) | 19.5 (0.10) |
| Total number of hospitalizations, between 2006 and 2016 |  |  |  |
| 0, % (SE) | 37.3 (0.12) | 1.8 (0.24) | 38.1 (0.13) |
| 1, % (SE) | 18.5 (0.10) | 12.4 (0.53) | 18.6 (0.10) |
| 2 to 5, % (SE) | 32.1 (0.12) | 45.8 (0.80) | 31.8 (0.12) |
| 6 and more, % (SE) | 12.1 (0.08) | 40.0 (0.78) | 11.5 (0.08) |

Notes: All percentages are weighted to the whole population of Canadians in 2006, using the CanCHEC cohort weight. Standard errors (SE) are calculated by bootstrapping. <sup>1</sup> Other includes individuals who were categorized under the "multiple minorities" or "Middle Easterns" groups in the Visible Minority question in Census. PESUH: people who experienced substance use harm during 2006-16; NPESUH: people who did not experienced substance use harm.

CIHI has previously highlighted a higher representation of substance use-related harm in rural areas, where rate ratios of rural age-standardized rate to urban age-standardized rates for hospitalization due to SUH among youth aged 10 to 24 in 2017-2018 were 1.5 and 1.7 for Saskatchewan and Canada respectively.^8^

In terms of ethnicity, there is a conspicuous overrepresentation of the Indigenous population within PESUH (41.6%), compared to their representation within the entire cohort (14.4%) and the NPESUH group (13.8%.) Conversely, while the cohort predominantly identified as white (82.0%), only 56.9% of PESUH and 82.6% of NPESUH did so. These observations align with a 2011 cohort study, in which hospitalization due to opioid poisoning (HOP) was notably higher among all Indigenous populations, especially for First Nations people living on reserve (5.6 times higher,) when compared to the non-Indigenous population.^17^ Additionally, a national study found that Indigenous students were significantly more likely to report poly-substance use compared to their white counterparts.^32^

More than one-third (41.5%) of PESUH were in the lowest income quintile at the study baseline. This is similar to CIHI’s report of the national level of substance use-related hospitalization in 2019.^33^ Another CIHI report describing the substance use-related hospitalizations among youth aged 10 to 24 also indicates similar numbers, with a rate ratio of 3.4 (in Saskatchewan) and 2.0 (in Canada) when comparing the age-standardized rates of SUH hospitalization between the lowest and highest neighbourhood income quintiles in 2017-2018.^8^ Carrière et al.’s study on HOP suggests similar trends, with higher crude rates of HOP (23.4 per 100,000) among people in the lowest income quintile compared to the HOP crude rate of 6.6 per 100,000 among people in the highest income quintile.^17^ Another national study on opioid-related harms also confirms our finding, reporting higher opioid-related harms among those in the lowest income quintile compared to the highest quintile, between 2000 and 2017.^5^

Similar to income, education was also inversely related to experiencing at least one SUH event over the study period. While 38.66% of the PESUH consists of people with educational attainment less than high school, this percentage for NPESUH and the cohort is comparatively lower, at 23.66% and 24% respectively. Similarly, Carrière et al. found that the age-standardized rate of HOP among people without a high school diploma was 2.1 times greater than the rates for those with a college diploma or university education below a bachelor’s degree.^17^

The PESUH group was more likely to be unemployed (9.31%) or not in labour force (36.64%) compared to the NPESUH group (3.78% and 24.43%, respectively.) We found that about one-third of PESUH in our cohort were employed in 2006, which is similar to the findings of a study on the socioeconomic profile of people who experienced an opioid overdose in B.C., where 33.8% of them were employed in the calendar year of their index overdose.^34^ The investigation of hospitalization due to opioid overdose also presents similar findings, with the highest rates of HOP among people who were unemployed (17.0 per 100,000.)^17^

Our findings indicate that PESUH are disproportionately more likely to work in “sales and services” (17.19%), “trades, transport and equipment operators and related occupations” (10.5%), and “occupations unique to processing, manufacturing and utilities” (2.5%), compared to their NPESUH counterparts (14.7%, 9.5%, and 1.9%, respectively.) Carrière et al.’s findings also indicate similar distribution, with the highest crude rates of HOP among people in “trade, transport equipment operators, and related occupations,” “sales and services,” and health, and the highest age-standardized rate among those in manufacturing and utility-related occupations.^17^ According to the BC cohort results, the majority of people who experienced an opioid overdose between 2014 and 2016 were primarily employed in construction industries (21.4%), followed by administrative and support, waste management and remediation services (12.2%), accommodation and food services (11.7%), retail trades (9.8%), and manufacturing (7.3%) industries.^34^ Since the BC cohort comparison was done using the North American Industry Classification System (NAICS), while we compared National Occupational Classification (NOC) categories, caution should be made in comparison of the BC findings with the SK results.

## Discussion

### Strengths and limitations of CanCHECs in constructing substance use cohorts

We compared our cohort with two others designed to study substance use harm in British Columbia^12^ and Saskatchewan.^13^ The comparison was primarily aimed at understanding the potential advantages and challenges of using CanCHECs, as in our study, versus using provincial data repositories, as was the approach in the BC^12^ and SK^13^ cohorts, in creating longitudinal cohorts for studying substance use harm.

A standout advantage of using CanCHECs for substance use research lies in its capacity to link socioeconomic and ethnocultural data of individuals to their health records. This allows an in-depth analysis of disparities in substance use harm based on factors such as income, education, occupation, language, self-identified ethnicity, including First Nations, Métis, and Inuit, and immigration status, among others.^15^ Such linkage is not possible with cohorts exclusively using provincial data repositories, as seen with the BC^12^ and SK^13^ cohorts.

CanCHEC is a national data source allowing for inter-provincial comparison, which enables researchers to conduct comparative policy analysis^35^ with regard to substance use countrywide. Moreover, the consistent linkage methodology used to create cohorts in CanCHEC provides the opportunity to examine trends over time both within and between CanCHEC cycles.^15^ CanCHEC also includes an annual historical postal code file from 1981 onward providing information on where the cohort members live year after year, which can be used to examine the impact of moving patterns on substance use harm. It should also be noted that CanCHEC administrative data updates continuously, enabling a prolonged follow-up of the individuals through time.

Nevertheless, there are limitations in using CanCHEC to investigate substance use harm. Notable, we are not able to identify substance use harms that did not result in hospitalization or death, using only CanCHEC. This is because of the absence of certain databases, such as pharmacy, community-based services, clinics, and physician billing data. These databases are usually accessible through provincial repositories.

Moreover, since CanCHEC excludes the institutional population^2^ (e.g., those living in nursing homes, penitentiaries, group homes) at baseline,^15^ the created cohort is representative of the non-institutional population living in Canada at the time of the census. This makes the cohort population younger and healthier than the Canadian population. Substance use harm events occurring in correctional settings and following release are well documented and particularly prevalent immediately following release,^36–45^ when many individuals are without an address or are residing in a group home so are not likely to be included with CanCHEC data. A recent scoping review also suggests that prescription misuse is a growing concern among older adults in Canada,^46^ suggesting that harm events in nursing homes may also be missed in the CanCHEC data. Group homes, or sober-living houses, also offer a supportive living environment for many people seeking abstinence-based recovery or treatment options.^47,48^ However, a return to use is common for almost half of the people engaging in recovery programs for substance use,^49^ and it is unlikely related harm events are captured by CanCHEC in these settings. Including institutional populations would increase the total number and frequency of harm events identified.

In general, census data quality reports indicate that a small proportion of Canadians is missed in any given census.^15^ These people are more likely to be young, mobile, low-income, homeless, or Indigenous.^15^ Given that these groups of the population are more likely to have experienced substance use harm,^5,17,50,51^ related estimates produced by CanCHEC are likely to be underestimated compared to the Canadian population. Therefore, considerable caution must be used when making inferences from CanCHEC, especially when generalizing to the broader population.

Nonetheless, although some populations are less likely to be fully captured by CanCHEC, there are others it is likely to capture more fully. For instance, it is likely to excel at portraying substance use behaviours amongst the middle class,^52^ which tends to be more likely to be employed and have higher education. Therefore, CanCHEC provides a great opportunity to investigate the characteristics of people who are outside the typical focus of substance use literature. A notable potential opportunity provided by CanCHEC is that it can also track the health outcomes of individuals who were teenagers or younger during a given census, providing potential insights into substance use and social mobility in adolescents and young adulthood.

### Increase in PESUH rate in 2016

The increase seen in the rates of PESUH in the final year of this study might be in part attributed to evolving national and provincial policies on substance use. For instance, in 2013, Dr. Fern Stockdale Winder’s appointment as a Commissioner resulted in the *Working Together for Change: A 10-Year Mental Health and Addictions Action Plan for Saskatchewan* report by December 2014.^53^ This report paved the way for increased mental health and substance use service awareness and destigmatization, with the recommendations being incorporated into various ministries’ agendas for a more comprehensive approach to mental health and substance use challenges.^54^ Nevertheless, further research should seek the underlying factors driving this trend.

### Study limitations

Our cohort of Saskatchewanians might inadvertently include individuals with hospitalization or mortality records over those without. This is due to our approach regarding identifying Saskatchewanians, where in addition to the census, we used DAD (2006-2016) and CVSD (2006-2016) data to find people who had an indication of residency in Saskatchewan.

Although the linkage error in CanCHEC is minimal, it is present,^15^ which could result in either an underestimation or overestimation of mortality and hospitalization rates attributed to SUH. Additionally, it is important to note that sociodemographic characteristics are only collected at the time of census, potentially not capturing shifts in an individual’s socioeconomic status throughout their life.

## Conclusion

The primary aim of this study was to use CanCHEC databases to create, describe, and validate a cohort of Saskatchewan residents focusing on substance use. We validated our cohort by comparing our descriptive findings with those from other Canadian studies on substance use. The Second objective was to underscore the potential CanCHEC holds in studying substance use, especially by bridging the gap in data concerning the linkage of social determinants of health and administrative health data. We hope this study can serve as a precursor to many more that use CanCHEC to examine the social determinants of substance use harm events using linked social and health administrative data. Future studies could focus on small-area analysis and multivariate analyses to understand the geographic distribution and social drivers of substance use harm in the province. Furthermore, researchers can adopt the STATA do-file developed by our research team to build their own cohorts using CanCHEC and investigate their research questions related to substance use (See Appendix E.)

Considering the findings of the current study, there is a clear need for research on possible changes in recording substance use hospitalizations by ICD codes in Canada. Understanding these changes is crucial in interpreting the rates of substance use harm. Further, policy impacts on substance use-related stigma and help-seeking behaviours warrant exploration, given their profound influence on substance use harm events statistics.

## Supporting information

Substance Use Cohort - Appendices A to D

Substance Use Cohort - Appendix E - STATA Do-File

## Author Contributions

AM conducted the data analysis and prepared the first draft of the article. AM and CP designed the study and directed its implementation, including quality assurance and control. CP supervised the data analysis. CP and FB reviewed, edited, and finalized the text. CP and FB provided the overall guidance and funding for the research project. All authors approved the final version of the manuscript.

## Acknowledgments

We are grateful to the Saskatchewan Research Data Centre analyst, Ruben Mercado, for all the help he provided in accessing data and vetting the results. We also thank Donica Janzen, a Senior Researcher at the Health Quality Council, and Raadiya Malam, a researcher and policy analyst at the Canadian Substance Use Costs and Harms (CSUCH), for the valuable information on substance use case-finding algorithm. The authors thank Carol Spencer, our patient family partner, for taking the time to give our research team excellent feedback and insightful comments. We also thank Daniel Yupanqui, Bong Soo Kim, and Shubrandu Sanjoy for their helpful comments throughout the project.

## Ethics Declaration

This study (REB-22-20) was deemed exempt by the University of Saskatchewan Research Ethics Board.

## Data Availability

All data used in this study is for public use and can be accessed through university libraries and the Data Liberation Initiative.

## Code Availability

Codes are available as a supplementary file to this working paper.

## Footnotes

1 In Census, “White” is one of the response categories in the population group question. where people are asked to respond to in the Visible Minority question.^30^ This is similar to what CIHI proposes in the “Guidance on the Use of Standards for Race-Based and Indigenous Identity Data Collection and Health Reporting in Canada” report. ^31^

2 CancCHEC considers individuals institutionalized if they do not have any other residency address in Canada and they have been in an institution for not less than six months.

