## Supplementary material for "Creating an 11-year longitudinal substance use harm cohort from linked health and census data to analyze social drivers of health": Substance Use Cohort - Appendices A to D

**Appendix A.** CanCHEC 2006 Data Structure.

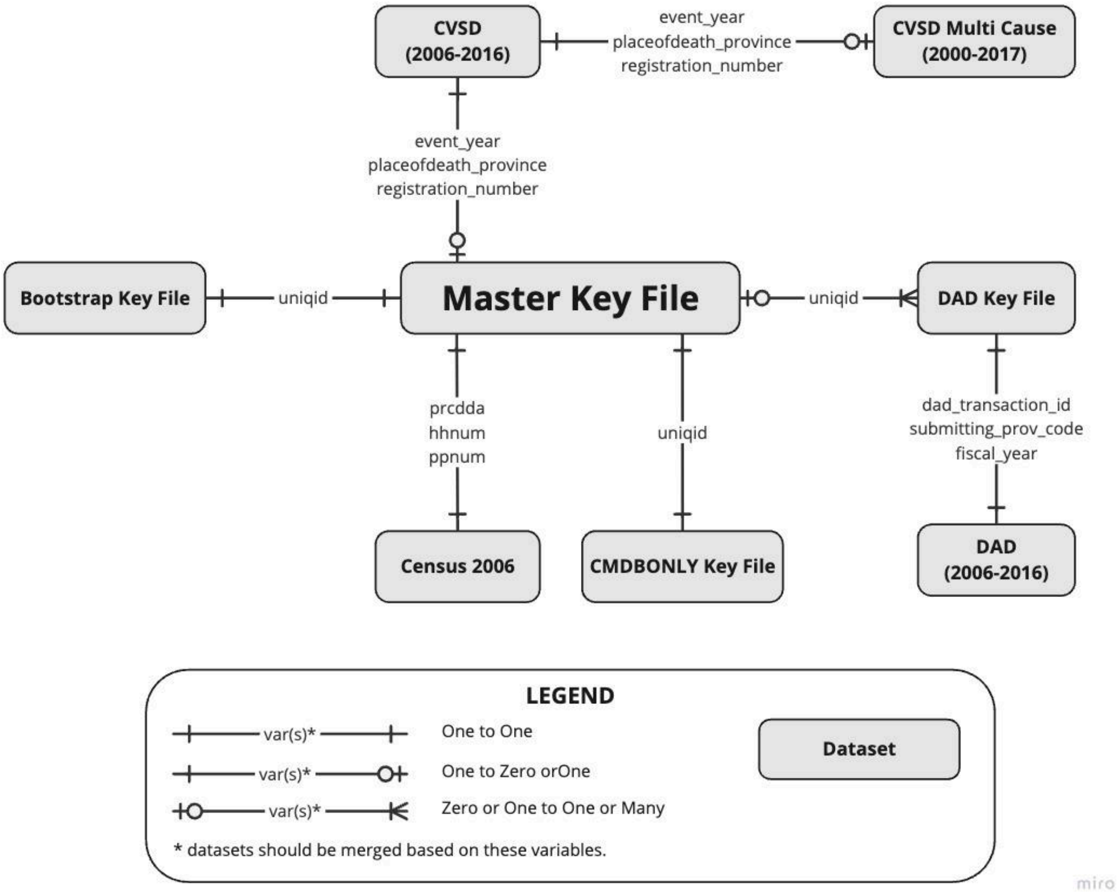


Note: This figure illustrates the relationship between different datasets available within CanCHEC 2006. The datasets shown in this figure only include those used in the current study and not all CanCHEC 2006 datasets. Data structures of different cycles of CanCHEC are different, and this diagram solely represents the cycle of 2006.

**Appendix B:** Substance use case finding algorithm.

| Database | Substance use category | Substance use case finding algorithm[1] | |
|---|---|---|---|
| | | **Diagnosis ICD-10-CA codes** | **Diagnosis type** |
| **DAD** | Alcohol | F10.–, T51.–, E24.4, G31.2, G62.1, G72.1, I42.6, K29.2, K70.–, K85.2, K86.0, O35.4–, O99.3–[2], Q86.0, R78.0, X45, X65, Y15, P04.3 | (M), (1), (2), (W), (X), (Y) or (9) |
| | Opioids | F11.–, T40.0, T40.1, T40.2–, T40.3, T40.4–, T40.6 O99.3–[2], P96.1 | (M), (1), (2), (W), (X), (Y) or (9) |
| | Cannabis | F12.–, T40.7, O99.3–[2] | (M), (1), (2), (W), (X), (Y) or (9) |
| | Other CNS depressants | F13.–, T42.3, T42.4, T42.6, T42.7, O99.3–[2] | (M), (1), (2), (W), (X), (Y) or (9) |
| | Cocaine | F14.–, T40.5, O99.3–[2] | (M), (1), (2), (W), (X), (Y) or (9) |
| | Other CNS stimulants | F15.–, T43.6, O99.3–[2] | (M), (1), (2), (W), (X), (Y) or (9) |
| | Unknown and multiple substances | F19.–, T43.8, T43.9, O99.3–[2], X41, X42, X61, X62, Y11, Y12[3], O35.5 | (M), (1), (2), (W), (X), (Y) or (9) |
| | Other substances[4] | F16.–, F18.–, F55, T40.8, T40.9, O99.3–[2] | (M), (1), (2), (W), (X), (Y) or (9) |
| | | **Underlying cause of death ICD-10-CA codes** | **Multiple cause of death ICD-10-CA codes** |
| **CVSD** | Alcohol | F10.–, E24.4, G31.2, G62.1, G72.1, I42.6, K29.2, K70.–, K85.2, K86.0, O35.4–, Q86.0, R78.0, X45, X65, Y15, P04.3 | -- |
| | | X40-X44, X60-X64, X85, Y10-Y14 | At least one comorbidity is T51.–. |
| | Opioids | F11.–, P96.1 | -- |
| | | X41-X44, X61-X64, X85, Y11-Y14 | At least one comorbidity is T40.0 (Opium), T40.1 (Heroin), T40.2 (Natural and semisynthetic opioids), T40.3 (Methadone), T40.4 (Synthetic opioids, other than methadone), or T40.6 (Other and unspecified narcotics). |
| | Cannabis | F12.– | -- |
| | | X40-X44, X60-X64, X85, Y10-Y14 | At least one comorbidity is T40.7. |
| | Other CNS depressants | F13.– | -- |
| | | X40-X44, X60-X64, X85, Y10-Y14 | At least one comorbidity is T42.3, T42.4, T42.6, T42.7. |
| | Cocaine | F14.– | -- |
| | | X40-X44, X60-X64, X85, Y10-Y14 | At least one comorbidity is T40.5 |
| | Other CNS stimulants | F15.– | -- |
| | | X40-X44, X60-X64, X85 Y10-Y14 | At least one comorbidity is T43.6. |
| | Unknown and multiple substances | F19.–, X41, X42, X61, X62, Y11, Y12, O35.5 | -- |
| | | X40-X44, X60-X64, X85, Y10-Y14 | At least one comorbidity is T43.8, T43.9. |
| | Other substances | F16.–, F18.–, F55 | -- |
| | | X40-X44, X60-X64, X85, Y10-Y14 | At least one comorbidity is T40.8, T40.9. |

Notes: [1] Conditions listed in both columns should be met in order for an event to be considered a SUH event. [2] Include only if F10–16 or F18–19 as diagnosis type (3) is in the same abstract. [3] Include X41, X61 and Y11 if neither T42.– nor T43.– are in the same abstract; include X42, X62 and Y12 if T40.– is not in the same abstract. [4] Includes hallucinogens, solvents, and abuse of non-dependence-producing substances. DAD: discharge abstract database; CVSD: Canadian vital statistics death database; ICD: international classification of diseases; CNS: central nervous system.

**Appendix C:** List of variables in the final derrived 2006 CanCHEC cohort.

| Name of variable in original dataset[1] | Final variable name | Variable label | Notes |
|---|---|---|---|
| uniqid | uniqid | unique ID (CanCHEC 2006) | Used in linking CanCHEC components based on Appendix A. |
| prcdda | da_06 | dissemination area (CanCHEC 2006) | Used in linking CanCHEC components based on Appendix A. |
| hhnum | hhnum | key for household table (CanCHEC 2006) | Used in linking CanCHEC components based on Appendix A. |
| ppnum | ppnum | key for person table (CanCHEC 2006) | Used in linking CanCHEC components based on Appendix A. |
| pp_id | pp_id | alternative key for census person table (CanCHEC 2006) | Used in linking CanCHEC components based on Appendix A. |
| event_year | year_death | year when death occurred (CanCHEC 2006) | Used in linking CanCHEC components based on Appendix A. |
| placeofdeath_province | placeofdeath_province | province where death occurred (CanCHEC 2006) | Used in linking CanCHEC components based on Appendix A. |
| registration_number | registration_number | death registration number (CanCHEC 2006) | Used in linking CanCHEC components based on Appendix A. |
| canchecw2 | w_cohort | CanCHEC 2006 weight (CanCHEC 2006) | CanCHEC 2006 weight. |
| dad_transaction_id | dad_transaction_id | DAD transaction ID (CanCHEC 2006) | Used in linking CanCHEC components based on Appendix A. |
| submitting_prov_code | submitting_prov_code | submitting province code (DAD) | Used in linking CanCHEC components based on Appendix A. |
| inst_code | inst_code | institution number (DAD) | Code assigned to a reporting facility by a provincial/ territorial ministry of health identifying the facility and the level of care of the data submitted in DAD. |
| fiscal_year | year_hosp | fiscal year of hospitalization (DAD) | The fiscal year of hospitalization, in DAD. |
| health_card_prov_code | hcard_pr | province issuing the health card (DAD) | Province issuing the health card of the patients, recorded in DAD. |
| patient_postal_code | resdn_pcode_dad | patient postal code (DAD) | Patient postal code recorded in DAD. |
| *derived variable*[2] | age_hosp | age at the time of hospitalization (DAD) | Age at the time of hospitalization in year, as recorded in DAD. This derived variable is based on the age_units and age_code variables in DAD. |
| gender_code | sex_hosp | sex of patient (DAD) | Sex of patiend recorded in the "gender_code" variable in DAD. |
| admission_date | admission_date | admission date (DAD) | Admission date in DAD. |
| discharge_date | discharge_date | discharge date (DAD) | Discharge date in DAD. |
| diag_cluster_`i' | diag_cluster_`i' | diagnosis cluster `i' (DAD) | This is a set of 25 variables, referring to the diagnosis cluster of an abstract in DAD. Diagnosis cluster is an alphabetic character which indicates association between diagnoses. |
| diag_prefix_`i' | diag_prefix_`i' | diagnosis prefix `i' (DAD) | This is a set of 25 variables, referring to the diagnosis prefix of an abstract in DAD. Diagnosis prefix is a one-character-long code which provides additional information on the ICD code to which it is assigned. |
| diag_code_`i' | diag_code_`i' | diagnosis code `i' (DAD) | This is a set of 25 variables, referring to the ICD-10 codes of the diagnosis for a patient record in DAD. |
| diag_type_`i' | diag_type_`i' | diagnosis type `i' (DAD) | This is a set of 25 variables, referring to the diagnosis type of an abstract in DAD. Diagnosis type is an alpha or numeric character used to describe the significance of a diagnosis or condition for a patient record in the DAD. |
| total_los_days | total_los_days | total length of stay (DAD) | This is the total length of stay as recorded in DAD. |
| acute_los_days | acute_los_days | acute length of stay (DAD) | This is the acute length of stay in hospital as recorded in DAD. |
| alc_los_days | alc_los_days | alternative level of care length of stay (DAD) | This is the alternative level of care length of stay in hospital, as recorded in DAD. |
| sex | sex_death | sex of deceased (CVSD) | This variable indicates the sex of deceased as available in CVSD. |
| *derived variable* | age_death | age of deceased (CVSD) | This is the age of deceased in year. This variable is derived form the age_value and age_code variables in CVSD. |
| residence_province_3digit | resdn_pr_cvsd | usual residence of deceased: province (CVSD) | This is the usual residence of deceased (province) available in CVSD. |
| residence_postalcode | resdn_pcode_cvsd | usual residence of deceased: postal code (CVSD) | This is the usual residence of deceased (postal code) available in CVSD. |
| death_cause_4digits | icd_underly_cause | ICD for underlying cause of death (CVSD) | This variable contains the ICD-10 codes of the underlying cause of death available in CVSD. |
| ra_mc_`i' | icd_contr_cause_`i' | ICD for contributing cause of death `i' (CVSD) | This is a set of 20 variables that refer to the ICD-10 codes of the contributing causes of death. These variables are available in the Multi Cause datasets associated with CVSD. |
| age | age_cens | age (census 2006) | Refers to the age at last birthday (as of the census reference date, May 16, 2006). This variable is derived from Date of birth. |
| cma | cma_cens | census metropolitan area or census agglomeration of current residence (2006) | Area consisting of one or more neighbouring municipalities situated around a major urban core. A census metropolitan area must have a total population of at least 100,000 of which 50,000 or more live in the urban core. A census agglomeration must have an urban core population of at least 10,000. |
| sex | sex_cens | sex (census 2006) | Refers to the gender of the respondent in Census 2006. |
| *derived variable* | hhatinc_adj | household after-tax income adjusted for household size (census 2006) | The after-tax income of a household is the sum of the after-tax incomes of all members of that household, adjusted for the household size. This variable is derived fromn the household after-tax income (hhinc_at) and household size (nunits) variables in Census 2006. |
| fsaname | fsa_cens | forward sortation area (census 2006) | The first three characters of the postal code identify the forward sortation area (FSA). FSAs are associated with a postal facility from which mail delivery originates. The average number of households served by an FSA is approximately 8,000, but the number can range from zero to more than 60,000 households. |

| | | | |
|---|---|---|---|
| pcd | cd_cens | census division of current residence (census 2006) | This is the census division of current residence in census 2006. |
| pcsd | csd_cens | census subdivision of current residence (census 2006) | This is the census subdivision of current residence in Census 2006. |
| pop | pop_csd | population size group of current census subdivision of residence (census 2006) | This is the population size group of current census subdivision of residence in Census 2006. |
| pr | pr_cens | province or territory of current residence (census 2006) | This is the province variable in Census 2006, referring to the province or territory of current residence of the respondent. |
| ruindfg | rufg | rural, urban classification (census 2006) | This is "ruindfg" variable in Census 2006, referring to the 2006 rural or urban classification of the CSD where the person usually resided on May 16, 2005, one year prior to Census Day. |
| rusize | rusize | rural/urban size code (census 2006) | This is the rural/urban size code variable in Census 2006. |
| compw2 | w_cens | composite weight (perswt + occwtp -1) (census 2006) | This is the Composite Weight (PERSWT + OCCWTP-1) variable in Census (compw2). |
| *derived variable* | edu | education (census 2006) | This derived variable is based on "Highest certificate, diploma or degree" variable (hcdd) in Census 2006. |
| nocsbrd | occ | labour market activities : occupation broad categories (census 2006) | This is the Broad National Occupational Classification variable in Census 2006. |
| *derived variable* | race | ethnicity (census 2006) | This derived variable is based on the Visible minority variable (dvismin) in Census 2006. |
| *derived variable* | emp | employment status (census 2006) | This derived variable is based on the Labour force activity variable (lf71) in Census 2006. |
| *derived variable* | resdn_sk_dad_p | resident of Saskatchewan based on DAD | This variable indicates that if a cohort member was identified as a resident of Saskatchewan based on DAD. |
| *derived variable* | resdn_sk_cvsd_p | resident of Saskatchewan based on CVSD | This variable indicates that if a cohort member was identified as a resident of Saskatchewan based on CVSD. |
| *derived variable* | resdn_sk_cens_p | resident of Saskatchewan based on census 2006 | This variable indicates that if a cohort member was identified as a resident of Saskatchewan based on census 2006. |
| *derived variable* | resdn_sk_p | person residing in SK at some point between 2006 and 2016 | This variable indicates that if a cohort member was identified as a person residing in Saskatchewan at some point between 2006 and 2016, based on DAD or CVSD or Census 2006. |
| *derived variable* | hosp_e | hospitalization event | This variable indicates a hospitalization event due to any reason. |
| *derived variable* | hosp_p | hospitalized person | This variable indicates if a person was hospitalized at least once, between 2006 and 2016, due to any reason. |
| *derived variable* | death_e | death event | This variable indicates a death event. |
| *derived variable* | death_p | deceased person | This variable indicates if a person died between 2006 and 2016. |
| *derived variable* | hosp_alc_e | hospitalization event due to alcohol | This variable indicates a hospitalization event due to alcohol. |
| *derived variable* | hosp_opi_e | hospitalization event due to opioid | This variable indicates a hospitalization event due to opioid. |
| *derived variable* | hosp_can_e | hospitalization event due to cannabis | This variable indicates a hospitalization event due to cannabis. |
| *derived variable* | hosp_cnsdep_e | hospitalization event due to other CNS depressants | This variable indicates a hospitalization event due to other CNS depressants. |
| *derived variable* | hosp_coc_e | hospitalization event due to cocaine | This variable indicates a hospitalization event due to cocaine. |
| *derived variable* | hosp_cnsstim_e | hospitalization event due to other CNS stimulants | This variable indicates a hospitalization event due to other CNS stimulants. |
| *derived variable* | hosp_unkmult_e | hospitalization event due to unknown and multiple substances | This variable indicates a hospitalization event due to unknown and multiple substances. |
| *derived variable* | hosp_other_e | hospitalization event due to other substances | This variable indicates a hospitalization event due to other substances. |
| *derived variable* | hosp_su_e | hospitalization event due to substance use | This variable indicates a hospitalization event due to substance use. |
| *derived variable* | hosp_alc_p | person hospitalized due to alcohol | This variable indicates if a person was hospitalized at least once due to alcohol, between 2006 and 2016. |
| *derived variable* | hosp_opi_p | person hospitalized due to opioids | This variable indicates if a person was hospitalized at least once due to opioids, between 2006 and 2016. |
| *derived variable* | hosp_can_p | person hospitalized due to cannabis | This variable indicates if a person was hospitalized at least once due to cannabis, between 2006 and 2016. |
| *derived variable* | hosp_cnsdep_p | person hospitalized due to other CNS depressants | This variable indicates if a person was hospitalized at least once due to other CNS depressants, between 2006 and 2016. |
| *derived variable* | hosp_coc_p | person hospitalized due to cocaine | This variable indicates if a person was hospitalized at least once due to cocaine, between 2006 and 2016. |
| *derived variable* | hosp_cnsstim_p | person hospitalized due to other CNS stimulants | This variable indicates if a person was hospitalized at least once due to other CNS stimulants, between 2006 and 2016. |
| *derived variable* | hosp_unkmult_p | person hospitalized due to unknown and multiple substances | This variable indicates if a person was hospitalized at least once due to unknown and multiple substances, between 2006 and 2016. |
| *derived variable* | hosp_other_p | person hospitalized due to other substances | This variable indicates if a person was hospitalized at least once due to other substances, between 2006 and 2016. |
| *derived variable* | hosp_su_p | person hospitalized due to substance use | This variable indicates if a person was hospitalized at least once due to substance use, between 2006 and 2016. |
| *derived variable* | death_alc_e | death event due to alcohol | This variable indicates death event due to alcohol. |
| *derived variable* | death_opi_e | death event due to opioids | This variable indicates death event due to opioids. |
| *derived variable* | death_can_e | death event due to cannabis | This variable indicates death event due to cannabis. |
| *derived variable* | death_cnsdep_e | death event due to other CNS depressants | This variable indicates death event due to other CNS depressants. |
| *derived variable* | death_coc_e | death event due to cocaine | This variable indicates death event due to cocaine. |

| | | | |
|---|---|---|---|
| *derived variable* | death_cnsstim_e | death event due to other CNS stimulants | This variable indicates death event due to other CNS stimulants. |
| *derived variable* | death_unkmult_e | death event due to unknown and multiple substances | This variable indicates death event due to unknown and multiple substances. |
| *derived variable* | death_other_e | death event due to other substances | This variable indicates death event due to other substances. |
| *derived variable* | death_su_e | death event due to substance use | This variable indicates death event due to substance use. |
| *derived variable* | death_alc_p | person died due to alcohol | This vairable indicates a person who died due to alcohol, between 2006 and 2016. |
| *derived variable* | death_opi_p | person died due to opioids | This vairable indicates a person who died due to opioids, between 2006 and 2016. |
| *derived variable* | death_can_p | person died due to cannabis | This vairable indicates a person who died due to cannabis, between 2006 and 2016. |
| *derived variable* | death_cnsdep_p | person died due to other CNS depressants | This vairable indicates a person who died due to other CNS depressants, between 2006 and 2016. |
| *derived variable* | death_coc_p | person died due to cocaine | This vairable indicates a person who died due to cocaine, between 2006 and 2016. |
| *derived variable* | death_cnsstim_p | person died due to other CNS stimulants | This vairable indicates a person who died due to other CNS stimulants, between 2006 and 2016. |
| *derived variable* | death_unkmult_p | person died due to unknown and multiple substances | This vairable indicates a person who died due to unknown and multiple substances, between 2006 and 2016. |
| *derived variable* | death_other_p | person died due to other substances | This variable indicates a person who died due to other substances, between 2006 and 2016. |
| *derived variable* | death_su_p | person died due to substance use | person died due to substance use |
| *derived variable* | pesuh | person who experienced substance use harm | Person who experienced substance use harm between 2006 and 2016. Substance use harm is defined as hospitalization or death due to substance use. |

Notes: [1] All variables are listed in lower case, independet of how they are saved in the original dataset. [2] A *derived variable* is a variable that is generated or recoded using variables in CanCHEC 2006. DAD: discharge abstract database; CVSD: Canadian vital statistics death database; CanCHEC: Canadian census health and environment cohorts; CNS: central nervous system; CSD: census subdivision; ICD: international classification of diseases.

**Appendix D:** Rate of people who experienced susbtance use harm per 100,000 in Saskatchewan weighted to CanCHEC 2006.

| | Substance use harm category, rate per 100,000 (SE) | | | | | | | |
|---|---|---|---|---|---|---|---|---|
| Year | Alcohol | Opioids | Cannabis | Other CNS depressants | Cocaine | Other CNS stimulants | Unknown/ multiple substances | Other substances[1] |
| 2006 | 170.7 (9.78) | 40.5 (4.99) | 39.5 (4.93) | 43.5 (5.28) | 34.0 (4.25) | 29.0 (4.10) | 54.1 (5.12) | 27.0 (3.80) |
| 2007 | 179.9 (9.60) | 47.7 (4.90) | 52.3 (5.94) | 46.7 (4.96) | 44.2 (4.96) | 31.5 (3.84) | 69.3 (6.49) | 31.2 (3.95) |
| 2008 | 202.9 (10.76) | 46.1 (4.95) | 47.1 (4.81) | 49.6 (5.47) | 49.1 (5.46) | 35.7 (4.44) | 71.9 (6.35) | 30.9 (4.13) |
| 2009 | 215.6 (11.45) | 57.6 (5.88) | 56.6 (5.71) | 45.9 (5.14) | 54.5 (6.52) | 35.9 (4.73) | 68.8 (6.39) | 33.1 (4.55) |
| 2010 | 198.8 (10.55) | 59.6 (6.19) | 52.9 (5.84) | 50.4 (5.51) | 45.2 (5.75) | 34.2 (4.84) | 62.2 (6.42) | 32.9 (4.76) |
| 2011 | 210.9 (10.69) | 60.6 (6.08) | 64.8 (6.98) | 53.9 (5.76) | 44.0 (5.14) | 33.9 (4.53) | 70.5 (6.96) | 31.6 (4.49) |
| 2012 | 223.2 (12.18) | 62.2 (6.27) | 61.7 (6.58) | 55.4 (6.07) | 44.4 (5.33) | 34.7 (4.76) | 73.2 (6.16) | 33.5 (4.68) |
| 2013 | 209.5 (12.03) | 58.0 (6.48) | 63.3 (6.32) | 49.1 (6.04) | 42.2 (5.45) | 39.7 (5.39) | 72.8 (6.76) | 33.2 (5.04) |
| 2014 | 231.7 (11.43) | 67.6 (6.64) | 74.0 (7.28) | 49.5 (5.41) | 45.3 (5.54) | 43.3 (5.32) | 68.2 (6.81) | 28.2 (4.24) |
| 2015 | 224.2 (12.01) | 78.5 (7.21) | 75.3 (6.32) | 42.5 (4.92) | 44.6 (5.20) | 49.0 (5.92) | 79.6 (7.25) | 22.6 (3.63) |
| 2016 | 265.1 (12.82) | 91.8 (8.24) | 124.9 (8.75) | 70.1 (7.17) | 65.2 (6.94) | 76.3 (7.52) | 81.5 (7.54) | 39.7 (5.58) |

Notes: [1] Other substances include hallucinogens, solvents, abuse of non-dependence-producing substances.
