## Supplementary material for "Creating an 11-year longitudinal substance use harm cohort from linked health and census data to analyze social drivers of health": Substance Use Cohort - Appendix E - STATA Do-File: Substance Use Cohort - Appendix E .do

```
*1234567890123456789012345678901234567890123456789012345678901234567890123456789
*
*
* 2021 SHRF - Substance Use Harm (SUH) Project                              ****
* ============================================================================ *
* Summary: This do-file creates national and provincial (Saskatchewan) cohorts
*   of respondents to the 2006 Census (long-form). The cohort follows 
*   hospitalization and mortality information of the respondents from 2006 to
*   2016. The final dataset includes variables that identify hospitalizations
*   and deaths due to substance use.
*
* Programmer: Anousheh Marouzi
*
* Start date: December 20th, 2022
* Last update: January 9th, 2024
* ============================================================================ *

* COPYRIGHT                                                                 ****
* ---------------------------------------------------------------------------- *
* Users of our code are kindly requested to acknowledge and cite our paper in 
* any publications or presentations where the code or its derivatives are 
* utilized.
* ---------------------------------------------------------------------------- *

* ABOUT CANCHEC                                                             ****
* ---------------------------------------------------------------------------- *
* CanCHEC is a set of key files that enables linking Census long-form to 
* hospitalization, emergency, mortality, and cancer registry, among other 
* databases. This linkage happens at the individual level. That is, the 
* socioeconomic and ethnocultural information of the Census respondents can be 
* linked to their health records, such as hospitalization. Databases used in 
* this do-file are listed below:
*
*   - Discharge Abstract Database (DAD)
*   - Canadian Vital Statistics Death Database (CVSD)
*   - Census 2006 (long-form)
*
* The health records, such as hospitalization, of these individuals can be
* tracked from years before the Census date followed to the years after 
* the Census date. For example, in CanCHEC 2006, we can link the information 
* of Census 2006 respondents to their hospitalization records, from 2000 to 
* 2016 (fiscal years.) 
* 
* Each of the CanCHEC's key files can be merged with a database based on a 
* specific set of variables. Below is a summary of how these key files work:
*
*   1. Master key file: This key file contains all individuals included in 
*      CanCHEC 2006. In this dataset, individuals are uniquely identified by 
*      the variable "UniqID". This is a person-based dataset, and its number 
*      of observations equals the number of individuals in CanCHEC 2006. The 
*      Master key file is a correspondence file between 2006 CanCHEC, 2006 
*      Census, and mortality captured by CVSD and the Canadian Mortality 
*      Database (CMDB). 
*
*   2. Bootstrap key file: This key file contains the CanCHEC 2006 sampling
*      weight (CanCHECW2) along with the bootstrap weights. The "CanCHECW2" 
*      is the Census 2006 weight variable adjusted to represent the CanCHEC  
*      cohort. The bootstrap key file can be linked to the Master key file  
*      using "uniqid" variable.
*
*   3. CMDBonly key file: This dataset is a copy of a subset of CMDB records 
*      for CanCHEC deaths that do not link to a death record on the CVSD.
*
*   4. DAD key file: Similar to DAD, the DAD key file is an event-based 
*      dataset, where each observation represents one hospitalization event. 
*      The DAD key file enables researchers to link DAD annual files to the 
*      Master key file. In that, DAD annual files are merged to DAD key file 
*      based on three variables: "dad_transaction_id", "submitting_prov_code", 
*      and "fiscal_year". The resulting dataset can then be linked to the 
*      Master key file using "uniqid" variable.
*
*  Appendix A in the manuscript illustrates the relationship between CanCHEC's
*  components that are used in this do-file.
* ---------------------------------------------------------------------------- *

* NAMING FILES                                                              ****
* ---------------------------------------------------------------------------- *
* We name datasets in this do-file according to a set of rules to facilitate
* understanding of the process a dataset goes through. This do-file does not do
* any analysis, and its only purpose is to manipulate and link different 
* datasets to create a data product that can be used in further research. 
* Thereby, each defined program opens a dataset, manipulates it, and saves it 
* by a meaningful name. 
*
* An overview of the structure of a dataset file's name is presented below: 
*
*    canchec`cycle'_`main info'_`geo'_`years covered'_`extra info'_`stage'
*
* The explanation for each section of this structure is as follows:
*
* 1. CanCHEC cycle as prefix: (Mandatory)
*    Every dataset in this do-file must have a prefix indicating which CanCHEC
*    cycle it is related to. The CanCHEC cycle used in this do-file is 2006.
*    Therefore, all datasets used in this do-file have the prefix of 
*    "canchec2006". There is only one exception, and that is the CVSD file. 
*    This is because raw CVSD files provided to the researchers in RDC are not 
*    manipulated for CanCHEC, meaning that CVSD is Statistics Canada's dataset 
*    standing on its own, covering the whole Canadian population. While the 
*    provided DAD datasets for CanCHEC users are manipulated in a way that only 
*    contains information about individuals in CanCHEC.
*
* 2. Main information included in the dataset: (Mandatory)
*    This section of the file name indicates the main information included in 
*    the dataset. This is usually the original name of the dataset, such as
*    "census" for Census (long-form), "dad" for Discharge Abstract Database, 
*    and "cvsd" for Canadian Vital Statistics Death Database.
*   
* 3. Geographic area covered by the dataset: (If Applicable)
*    Where applicable, the geographic area covered by the dataset is mentioned
*    here. To show that the dataset covers the whole of Canada, we use "nat" 
*    (for national), and we use abbreviations of the provinces to show datasets 
*    limited to a province(s), such as "sk" for Saskatchewan.
*     
* 4. Year(s) covered by the dataset: (Mandatory)
*    Every dataset must have this part indicating the year(s) covered by the 
*    dataset. For example, "2006", or "2006to2016".
*
* 5. Extra information about what exists in the dataset: (If Needed)
*    We add more information in the name of a dataset where we find it helpful
*    to communicate what exists in a dataset. Explanations for this extra
*    information are provided at the beginning of the program. 
*    Some examples include:
*      "_e"  --> indicating that this is an event-based dataset.
*      "_p"  --> indicating that this is a person-based dataset.
*      "_all_hospdeath" --> indicating that the dataset has dummy variables  
*        for all hospitalizations and deaths.
*      "_su_hosp" --> indicating that the dataset has dummy variables for 
*        substance use- (su) related hospitalizations
*
* 6. Stage of the dataset as suffix: (Mandatory)
*    Files in this do-file are classified into three main groups: Raw, Master, 
*    and Use. These are indicative of three main steps that a dataset usually
*    goes through in this do-file; A dataset matures from being a Raw file into  
*    a Master file and finally becomes a Use file. However, these are not the  
*    only three statuses of a dataset. Between the Raw and Master steps, we may  
*    save the dataset as a Reduced or Harmonized file. Following is a flow    
*    chart of how a dataset is processed in this do-file: 
*    
*             RAW --> (REDUCED) --> (HARM) --> MASTER --> USE 
*    
*    We add a suffix to the name of a dataset to show these stages. Below, we 
*    describe these 5 stages and provide the suffix in parentheses as they  
*    exist in the do-file:
*
*      a. Raw (_RAW): The first step in this do-file is to rename all raw  
*         datasets files according to our naming structure. This is done to    
*         have a consistent and meaningful name convention in our do-files 
*         across all our projects. We put the suffix "_RAW" at the end of the 
*         renamed raw files. These files are 100% similar to the original ones 
*         as provided in RDC.
*    
*      b. Reduced (_REDUCED): We add this suffix to datasets that we have 
*         dropped some of its variables or observations (reduced the number of 
*         variables or observations.) This step is usually done to speed up  
*         run times.
*     
*      c. Harmonized (_HARM): This suffix is added to datasets when its
*         variables are manipulated in a way to be harmonized in relation to
*         other datasets in the do-file. For example, variable sex is recorded
*         in different datasets to be consistent with how it is coded in the 
*         Census.
*     
*      d. Master (_MASTER): The Master stage of a dataset happens when general 
*         manipulations, such as reducing the number of variables and recoding
*         them, have been done, and the dataset is ready for more specialized
*         manipulation. A key feature of a Master file is that it usually 
*         contains the same number of rows as the raw file.
*    
*      e. Use (_USE): We add the suffix "USE" to a dataset when we have 
*         manipulated the Master dataset in a way that is ready for analysis.
*         This analysis can be descriptive, modeling, or any other type of 
*         analysis that produces the results of a research project. This step 
*         usually involves generating variables and dropping unnecessary 
*         observations.
* ---------------------------------------------------------------------------- *

* List of datasets in this do-file: 
* ---------------------------------------------------------------------------- *
*  Inputs:
*    1. keyfile_v1b_2006.dta
*    2. CanCHEC_bs_2006.dta
*    3. cmdbonly2006.dta
*    4. CanCHEC_2006_dadkey_f3_v1.dta
*    5. cen_2006_f1_v2.dta
*    6. cvsd`year'_f1_v2.dta                   for each year 2006-2010
*    7. vsd_death_2011_f1_v3.dta 
*    8. vsd_death_2012_f1_v2.dta
*    9. vsd_death_`year’_f1_v1.dta             for each year 2013-2015
*    10. vsd_sec_death_2016_f1_v1.dta
*    11. multiplecause`year'_f1_v2.dta         for each year 2006-2016
*    12. canchec_2006_dad_`year'_f3_v1.dta     for each year 2006-2016
*        
*  Outputs:
*    1. canchec2006_key_master_HARM
*    2. canchec2006_key_bs_HARM
*    3. canchec2006_key_cmdbonly_HARM
*    4. canchec2006_dad_`year'_REDUCED for each year 2006-2016
*    5. canchec2006_dad_2006to2016_MASTER
*    6. cvsd_`year'_REDUCED for each year 2006-2016
*    7. cvsd_2006to2016_HARM
*    8. cvsd_2006to2016_MASTER
*    9. census_long_2006_MASTER
*    10. canchec2006_cohort_can_06to16_e_USE
*    11. canchec2006_cohort_sk_06to16_e_USE
*    12. canchec2006_cohort_sk_06to16_all_hospdeath_e_MASTER
*    13. canchec2006_cohort_sk_06to16_all_hospdeath_su_hosp_e_MASTER
*    14. canchec2006_cohort_sk_06to16_all_hospdeath_su_hospdeath_e_USE
*    15. canchec2006_cohort_sk_06to16_all_hospdeath_su_hospdeath_e_FINAL
* ---------------------------------------------------------------------------- *

* TABLE OF CONTENTS                                                         ****
* ---------------------------------------------------------------------------- *
* 0. SETUP
* 1. RENAME RAW DATA FILES 
* 2. HARMONIZE ALL CANCHEC 2006 KEY FILES
* 3. PREPARE DAD (2006 TO 2016)                                             
* 4. PREPARE CVSD (2006 TO 2016)                                            
* 5. PREPARE CENSUS 2006 (LONG-FORM)                                        
* 6. CREATE NATIONAL COHORT
* 7. CREATE PROVINCIAL COHORT (SASKATCHEWAN)
* 8. IDENTIFY SUBSTANCE USE HARM EVENTS AND PESUH	
* A1. CONTROL
* ---------------------------------------------------------------------------- *

*  *Version 13                                                              ****
/* - - - - - - - - - - - - - - - - - - - - - - - - - - - - - - - - - - - - - - * 

   - This version tends to improve the readability of the codes in a way that
     is shareable with others. This version only contains the part of the code 
     where the cohort is created.
	 
* - - - - - - - - - - - - - - - - - - - - - - - - - - - - - - - - - - - - - - */

* *Empty Note                                                               ****
/* - - - - - - - - - - - - - - - - - - - - - - - - - - - - - - - - - - - - - - * 

  Here is a note that can be copied and pasted and used as a template.
   
* - - - - - - - - - - - - - - - - - - - - - - - - - - - - - - - - - - - - - - */

* 0. SETUP                                                                  ****
* ---------------------------------------------------------------------------- *
* Global setiings
set more off, permanently
clear all

* Name project
local projectname SHRF
local filename create_cohort

* Set directory
cd P:\Proj-10058_FFS-Plante_Sanjoy\WORK\Anousheh
	
* Set ado path
adopath + ".\ADO"
	
* Log
capture log close main
local date = subinstr(c(current_date)," "," ",.)
log using "./Log/`filename'_`date'.log", name(main) replace
	
* Set graph
set scheme s1color

* 1. MOVE AND RENAME RAW DATA FILES                                         ****
* ---------------------------------------------------------------------------- *
program define move_raw
  * Open the raw file of CanCHEC 2006 Master Keyfile to rename it
  use "S:\CanCHEC_CSERCan_2006\rdc_cdr\CanCHEC_CSERCan_2006_v2\data_donnees\stata\keyfile_v1b_2006.dta" ///
    , clear
	
  * Save the raw file according to our naming conventions
  save ".\Data\canchec2006_key_master_RAW.dta", replace
  
  * Open the raw file of CanCHEC 2006 Bootstrap Keyfile to rename it
  use "S:\CanCHEC_CSERCan_2006\rdc_cdr\CanCHEC_CSERCan_2006_v2\data_donnees\bootstrap\stata\CanCHEC_bs_2006.dta" ///
    , clear
	
  * Save the raw file according to our naming conventions
  save ".\Data\canchec2006_key_bs_RAW.dta", replace
  
  * Open the raw file of CanCHEC 2006 CMDBonly Keyfile to rename it
  use "S:\CanCHEC_CSERCan_2006\rdc_cdr\CanCHEC_CSERCan_2006_v2\data_donnees\stata\cmdbonly2006.dta" ///
    , clear
	
  * Save the raw file according to our naming conventions
  save ".\Data\canchec2006_key_cmdbonly_RAW.dta", replace
   
  * Open the raw files of CanCHEC 2006 DAD annual files (2006-2016) to rename it
  forvalues year = 2006/2016 { 
    use "S:\CanCHEC_CSERCan_2006_DAD\rdc_cdr\CanCHEC_2006_DAD_v1\data_donnees\data\DAD\stata_en\canchec_2006_dad_`year'_f3_v1.dta" ///
      , clear
	  
    * Save the raw file according to our naming conventions
    save ".\Data\canchec2006_dad_`year'_RAW.dta", replace
  }
  
  * Open the raw file of CanCHEC 2006 DAD Keyfile to rename it
  use "S:\CanCHEC_CSERCan_2006_DAD\rdc_cdr\CanCHEC_2006_DAD_v1\data_donnees\data\DAD_keys\stata_en\CanCHEC_2006_dadkey_f3_v1.dta" ///
    , clear
	
  * Save the raw file according to our naming conventions
  save ".\Data\canchec2006_key_dad_RAW.dta", replace
  
  * Open the raw files of CVSD annual files (2006-2010) to rename it
  forvalues year = 2006/2010 {
    use "S:\VSDD_SECD_AllYears\rdc_cdr\VSDD_SECD_19742011_v4\data_donnees\data\stata_en\cvsd`year'_f1_v2.dta" ///
      , clear
  
    * Save the raw file according to our naming conventions
    save ".\Data\cvsd_`year'_RAW.dta", replace
  }
  
  * Open the raw files of CVSD annual files (2011-2016) to rename it
  * Open CVSD 2011
  use "S:\VSDD_SECD_AllYears\rdc_cdr\VSDD_SECD_2011_v3\data_donnees\data\stata_en\vsd_death_2011_f1_v3.dta" ///
   , clear
	
  * Save and rename CVSD 2011
  save ".\Data\cvsd_2011_RAW.dta", replace
  
  * Open CVSD 2012	
  use"S:\VSDD_SECD_AllYears\rdc_cdr\VSDD_SECD_2012_v3\data_donnees\data\stata_en\vsd_death_2012_f1_v2.dta" ///
    , clear

  * Save and rename CVSD 2012
  save ".\Data\cvsd_2012_RAW.dta", replace
  
  * Open CVSD 2013	
  use "S:\VSDD_SECD_AllYears\rdc_cdr\VSDD_SECD_2013_v2\data_donnees\data\stata_en\vsd_death_2013_f1_v1.dta" ///
    , clear

  * Save and rename CVSD 2013
  save ".\Data\cvsd_2013_RAW.dta", replace
  
  * Open CVSD 2014	
  use "S:\VSDD_SECD_AllYears\rdc_cdr\VSDD_SECD_2014_v1\data_donnees\data\stata_en\vsd_death_2014_f1_v1.dta" ///
    , clear

  * Save and rename CVSD 2014
  save ".\Data\cvsd_2014_RAW.dta", replace
  
  * Open CVSD 2015	
  use "S:\VSDD_SECD_AllYears\rdc_cdr\VSDD_SECD_2015_v1\data_donnees\data\stata_en\vsd_death_2015_f1_v1.dta" ///
    , clear 

  * Save and rename CVSD 2015
  save ".\Data\cvsd_2015_RAW.dta", replace
  
  * Open CVSD 2016	
  use "S:\VSDD_SECD_AllYears\rdc_cdr\VSDD_SECD_2016_v1\data_donnees\data\stata_en\vsd_sec_death_2016_f1_v1.dta" ///
    , clear

  * Save and rename CVSD 2016
  save ".\Data\cvsd_2016_RAW.dta", replace
  
  * Open the raw files of CVSD Multi Cause file (2006-2016) to rename it
  forvalues year = 2006/2016 {
    use "S:\VSDD_SECD_AllYears\rdc_cdr\VSDD_SECD_MULTCAUS_20002017_v1\data_donnees\data\stata\multiplecause`year'_f1_v2.dta" ///
      , clear
  
    * Save the CVSD Multi Cause file according to our naming conventions
    save ".\Data\cvsd_multi_cause_`year'_RAW.dta", replace
  }
  
  * Open the raw file of Census 2006 (long-form) to rename it
  use "S:\CEN_REC_2006\rdc_cdr\CEN_REC_2006_v2\data_donnees\data\stata_en\cen_2006_f1_v2.dta" ///
    , clear
	
  * Save the Census file according to our naming conventions	
  save ".\Data\census_long_2006_RAW.dta", replace
end
	
* 2. HARMONIZE ALL CANCHEC 2006 KEY FILES                                   ****
* ---------------------------------------------------------------------------- *
* This Section harmonizes CanCHEC 2006 key files that are going to be used in 
* this do-file. This is done to change all variables to similar ones across  
* ALL DATASETS to enable a smooth merging of different files based on 
* different variables in Section 6. This step is necessary for creating the 
* cohort as we need the variables to be homogenous when we merge them 
* together.

* Out of the four CanCHEC keyfiles used in this do-file, we harmonize three
* of them, namely: Master Keyfile, Bootstrap Keyfile, and CMDBonly Keyfile.
* We did not need to harmonize the DAD key file. We used the raw DAD Keyfile, 
* as provided in Folder S in RDC, in Section 2.

program define harm_keyfile_master
  * Renames and recodes variables in CanCHEC Master Keyfile to be similar to 
  * what exists in other datasets in CanCHEC.
  
  * Open the raw CanCHEC 2006 Master key file 
  use ".\Data\canchec2006_key_master_RAW.dta", clear
	
  * Rename variables to lowercase to be similar to what we have in other 
  * datasets
  rename *, lower
  
  * Save as numeric if you can (this will also fix the missing values which are 
  * saved such as "    .")
  destring *, replace

  * Save
  save ".\Data\canchec2006_key_master_HARM.dta", replace
end

program define harm_keyfile_bs	
  * This program renames and recodes variables in the CanCHEC Bootstrap Keyfile  
  * to be similar to what exists in other datasets in CanCHEC. 
  
  * Open the raw CanCHEC 2006 bootstrap key file with two variables we need
  use UniqID CANCHECW2 using ".\Data\canchec2006_key_bs_RAW.dta", clear
  
  * Rename variables
  rename CANCHECW2 w_cohort
  rename UniqID uniqid
  
  * Save a smaller size of the bootstrap key file
  save ".\Data\canchec2006_key_bs_HARM.dta", replace
end

program define harm_keyfile_cmdbonly		
  * This program renames and recodes variables in the CanCHEC CMDBonly Keyfile 
  * to be similar to what exists in other datasets in CanCHEC. 
  
  * Open raw CanCHEC 2006 CMDBonly Keyfile
  use ".\Data\canchec2006_key_cmdbonly_RAW.dta", clear
 
  * Keep variables we need
  keep EVENT_YEAR PLACEOFDEATH_PROVINCE REGISTRATION_NUMBER SEX AGE RESPROV  ///
    POSTAL CAUSE PP_ID UniqID PRCDDA PpNum
 
  * Rename variables
  rename *, lower
  rename resprov 	resdn_pr_cvsd
  rename postal		resdn_pcode_cvsd
  rename cause 		icd_underly_cause
 
  * Generate age variable
  generate age_death = substr(age,3,2) if substr(age,1,2) == "50"
  
  * Dropping unnecessary characters from age variable (CMDBonly)            ****
  /* - - - - - - - - - - - - - - - - - - - - - - - - - - - - - - - - - - - - - *
     In the CMDBonly key file dataset, the age variable is saved as a string 
     starting with "50". For example, if a person died at 49 years of age, the 
     age variable is saved as "5049". Because of this, "50" was dropped from  
     the beginning of the age values to get the age of the deceased.
  * - - - - - - - - - - - - - - - - - - - - - - - - - - - - - - - - - - - - - */  
  
  * Generate sex variable
  generate sex_death = 2 if sex == "1"
  replace sex_death = 1 if sex == "2"
  
  * Changing values of sex variable in CVSD according to how it is coded in ****
  * Census   
  /* - - - - - - - - - - - - - - - - - - - - - - - - - - - - - - - - - - - - - *
     In CMDBonly Keyfile and CVSD, the sex variable is saved as "1" for males 
     and "2" for females. However, in Census 2006, female is coded as "1" and 
     male as 2. To be consistent across all datasets, the sex variables in CVSD  
     and CMDBonly key file are changed to be similar to the Census' sex 
     variable.
  * - - - - - - - - - - - - - - - - - - - - - - - - - - - - - - - - - - - - - */    
  
  * Drop the original sex and age variables
  drop sex age
  
  * Save as numeric if you can
  destring *, replace
  
  * Save		
  save ".\Data\canchec2006_key_cmdbonly_HARM.dta", replace			
end
		
* 3. REDUCE DAD (2006 TO 2016)                                              ****
* ---------------------------------------------------------------------------- *
program define reduce_dad
  * This program reduces the number of variables in DAD annual files and only 
  * keeps those that are more likely to be used in the final cohort. The  
  * reduced datasets are saved separately for each year.

  * The list of variables can be updated based on researchers' needs. This can
  * be done by consulting DAD User Guides available in RDC. Furthermore, 
  * researchers can change the period of time they want to follow 
  * hospitalization records by adding/eliminating years used in this program
  * (i.e., 2006 to 2016.)
  
  * Start a loop to reduce DAD files for each year 
  forvalues year = 2006/2016 { 
  
    * Open DAD raw annual files
    use ".\Data\canchec2006_dad_`year'_RAW.dta", clear
	
    * Keep variables we need
    keep dad_transaction_id submitting_prov_code inst_code fiscal_year       ///
      health_card_prov_code patient_postal_code admission_date               ///
      discharge_date age_code age_units gender_code total_los_days           ///
	  acute_los_days alc_los_days diag*         
		
    * Save as numeric if you can
    destring *, replace	
		
    * Save
    save ".\Data\canchec2006_dad_`year'_REDUCED.dta", replace	
  }
end 

* 4. APPEND DAD (2006 TO 2016)                                              ****
* ---------------------------------------------------------------------------- *
program define append_dad	
  * This program appends reduced DAD annual files (2006 to 2016) and merges the 
  * result with DAD keyfile. This merge enables linking DAD to the Master key 
  * file in Step 5.
 
  * Open 2006 reduced DAD dataset
  use ".\Data\canchec2006_dad_2006_REDUCED.dta", clear

  * Append years 2007 to 2016
  forvalues year = 2007/2016 {
    append using ".\Data\canchec2006_dad_`year'_REDUCED.dta"
  }
  
  * Merge with CanCHEC 2006 DAD key file to be able to merge with Master 
  * Keyfile in the Step 5
  merge 1:1 dad_transaction_id submitting_prov_code fiscal_year              ///
    using ".\Data\canchec2006_key_dad_RAW.dta",	nogen keep (3)  

  *  Merge results                                                          ****
  /* - - - - - - - - - - - - - - - - - - - - - - - - - - - - - - - - - - - - - *
   - MASTER FILE:
       The number of observations equals the number of abstracts recorded in  
       DAD from 2006 to 2016. This is the same as the number of hospitalizations 
       that happened to the CanCHEC 2006 cohort in these years.
     
   - USING FILE:
       The number of observations equals the number of abstracts recorded in 
       DAD from 2000 to 2016. This is the same as the number of hospitalizations
       that happened to the CanCHEC 2006 cohort in these years.
  	
   - MERGE RESULT:
       Matched = The number of observations in the Master File.
       
       Not matched from master = All observations are matched.
       
       Not matched from using = The number of hospitalization records happened 
         between the years 2000 and 2005.
  * - - - - - - - - - - - - - - - - - - - - - - - - - - - - - - - - - - - - - */
		
  * Recode age and only keep the derived variable
  replace age_units = 0 if age_code != "Y"		
  drop age_code
  
  *  Note on age variable in DAD                                            ****
  /* - - - - - - - - - - - - - - - - - - - - - - - - - - - - - - - - - - - - - *
     The age variable in DAD is represented by concatenating two pieces of
     information:
     
       1. Age Unit: is numeric values that measure the specified Age Code.
       2. Age Code: is the value denoting how the patient's age is measured. 
          For example, "Y" indicates that age is measured by year.
  
     We showed the age of people younger than a year old by zero.	   
  * - - - - - - - - - - - - - - - - - - - - - - - - - - - - - - - - - - - - - */    
	
  * Generate sex to numbers to be consistent across datasets
  generate sex_hosp = 0 if gender_code == "O"		
  replace sex_hosp = 2 if gender_code == "M"
  replace sex_hosp = 1 if gender_code == "F"
  
  * Drop the original sex variable
  drop gender_code
  
  * Rename variables
  rename fiscal_year            year_hosp
  rename health_card_prov_code 	hcard_pr
  rename patient_postal_code 	resdn_pcode_dad
  rename age_units              age_hosp			
  
  * Save as numeric if you can
  destring *, replace
    
  * Save
  save ".\Data\canchec2006_dad_2006to2016_MASTER.dta", replace	
	
  *  How to use the saved DAD Master file                                   ****
  /* - - - - - - - - - - - - - - - - - - - - - - - - - - - - - - - - - - - - - *
     The final DAD MASTER file can be used in Step 5 when we are compiling all 
     datasets to create the national cohort. This DAD MASTER file can be linked 
     to the Master Keyfile by uniqid, so that we include hospitalization  
     records in the final cohort.
  * - - - - - - - - - - - - - - - - - - - - - - - - - - - - - - - - - - - - - */	
end		
		
* 5. REDUCE CVSD (2006 TO 2016)                                             ****
* ---------------------------------------------------------------------------- *
program define reduce_cvsd
  * This program harmonizes CVSD from 2006 to 2016 and saves the results in 
  * separate file for each year.
  
  * Start a loop to reduce the number of variables in CVSD annual files
  forvalues year = 2006/2016{
	
    * Open raw data
    use ".\Data\cvsd_`year'_RAW.dta", clear
    
    * Rename and drop variable named "long" in 2006 dataset
    if inlist(`year', 2006, 2007, 2008, 2009, 2010) {
      rename long long_var
    }
	
    *  Issues with a variable named "long" in CVSD 2006 - 2010              ****
    /* - - - - - - - - - - - - - - - - - - - - - - - - - - - - - - - - - - - - *
       In CVSD 2006 to 2010, there is a variable named "long." Since "long" is
       a reserved keyword in STATA, the next line of codes, where we intend to
       rename all variables to lower case (i.e., rename *, lower), does not 
       work. This is because STATA cannot rename any variable to "long." 
	   
       To address this problem, we rename the variable named "long" to 
       "long_var", so that we can rename all variables to the lowercase 
       without any error.	
    * - - - - - - - - - - - - - - - - - - - - - - - - - - - - - - - - - - - - */   		

    * Rename variables to lowercase
    rename *, lower
	
    * Keep variables we need
    keep event_year placeofdeath_province registration_number sex age_code   ///
      age_value residence_province_3digit residence_postalcode               ///
      death_cause_4digits
	  
    * Save as numeric if you can
    destring *, replace
    	
    * Save reduced datasets
    save ".\Data\cvsd_`year'_REDUCED.dta", replace
  }
end

* 6. APPEND CVSD (2006 TO 2016)                                             ****
* ---------------------------------------------------------------------------- *
program define append_cvsd
  * This program appends harmonized CVSD datasets (2006 to 2016) resulting in  
  * the previous step.
	
  * Open the first harmonized CVSD dataset (2006)
  use ".\Data\cvsd_2006_REDUCED.dta", clear	
  
  * Append CVSD for years 2007-2016
  forvalues year = 2007/2016 {
    append using ".\Data\cvsd_`year'_REDUCED.dta" 
  }	
  
  * Recode the age variable and only keep the derived variable
  replace age_value = 0 if inlist(age_code,1,2,3,4)		
  drop age_code
  
  *  Note on age variables in CVSD                                          ****
  /* - - - - - - - - - - - - - - - - - - - - - - - - - - - - - - - - - - - - - *
     In CVSD, the age of death is represented by two variables:
     
     1. Age_value: Actual age of deceased reported in time units of either 
  	  minutes, hours, days, months, or years (as specified by the age 
          code.)
     2. Age_code: This variable indicates the unit of time measurement used to 
  	  report the age of the deceased (age value.)
  	   
  	     age_code | Measureing
  		 ---------+-----------
  		    1     | Minutes
  		    2     | Hourse
  		    3     | Days
  		    4     | Months
  		    5     | Years	
  		    9     | Unknown
  	 
     We showed the age of people deceased younger than one year old by zero.	   
  * - - - - - - - - - - - - - - - - - - - - - - - - - - - - - - - - - - - - - */   		
  
  * Generate sex variable so that it is similar to how it is coded in the Census
  generate sex_death = 0 if sex == 9
  replace sex_death = 1 if sex == 2
  replace sex_death = 2 if sex == 1

  * Drop the original sex variable
  drop sex
  
  *  Note on sex variable in CanCHEC                                        ****  
  /* - - - - - - - - - - - - - - - - - - - - - - - - - - - - - - - - - - - - - *
     It should be noted that while the sex variable in Census is coded as 1 for 
     female and 2 for male, this is the other way around in CVSD. So, we 
     changed the CVSD's sex variable to match what we have in the Census.
  * - - - - - - - - - - - - - - - - - - - - - - - - - - - - - - - - - - - - - */
 
  * Rename variables
  rename residence_province_3digit  resdn_pr_cvsd
  rename residence_postalcode       resdn_pcode_cvsd
  rename death_cause_4digits        icd_underly_cause
  rename age_value                  age_death	
  
  * Save as numeric if you can
  destring *, replace
  
  * Save appended dataset
  save ".\Data\cvsd_2006to2016_HARM.dta", replace		
end 

* 7. MERGE CVSD TO CVSD MULTI CAUSE (2006 TO 2016)                          ****
* ---------------------------------------------------------------------------- * 
program define merge_cvsd_multi
  * This program merges harmonized CVSD datasets with the CVSD Multiple Cause 
  * dataset.

  * The reason to do so is that in order to identify deaths due to substance 
  * use POISONING, we need to consider variables (i.e., contributing cause,)  
  * which only exists in the Multiple Cause dataset. 

  * The contributing cause information is included in a set of 20 variables 
  * named "ra_mc_`i'", where i is 1 to 20. These variables provide additional 
  * information for each death registered in the main CVSD files, which MUST  
  * be used to identify SPECIFIC deaths, such as deaths due to substance use 
  * poisoning. That is, the contributing cause variables are not useful to 
  * identifying EVERY TYPE OF DAETH. For example, we don't need to consider  
  * them when identifying deaths due to non-poisoning substance use causes. 

  * Open the first CVSD Multi Cause dataset (2006) to append to the other years
  use ".\Data\cvsd_multi_cause_2006_RAW.dta", clear
  
  * Append to CVSD Multi Cause datasets (2007-2016)
  forvalues year = 2007/2016 {
    append using ".\Data\cvsd_multi_cause_`year'_RAW.dta"
  }	
  
  * Rename variables to lowercase
  rename *, lower
  
  * Save as numeric if you can
  destring *, replace
  
  * Keep variables we need in the CVSD Multi Cause appended file (2006-2016)
  keep event_year placeofdeath_province registration_number ra_mc*

  *  How CVSD main file relates to Multi Cause file                         ****
  /* - - - - - - - - - - - - - - - - - - - - - - - - - - - - - - - - - - - - - *
     The number of observations recorded for the years 2006 to 2016 in the 
     appended Multi Cause file is less than the number of deaths recorded in 
     the main CVSD file for these years. This may be because of no existing 
     records for contributing causes of death for those observations missing 
     in the Multi Cause file.
  * - - - - - - - - - - - - - - - - - - - - - - - - - - - - - - - - - - - - - */
				
  * Merge with harmonized CVSD 2006 to 2016
  merge 1:1 event_year placeofdeath_province registration_number using       ///
    ".\Data\cvsd_2006to2016_HARM.dta", nogen keep(2 3)

  *  Merge results                                                          ****	
  /* - - - - - - - - - - - - - - - - - - - - - - - - - - - - - - - - - - - - - *
   - MASTER FILE:
       The number of observations in the Master File equals the number of   
       deaths in Multi Casue file from 2000 to 2016.
  	 
   - USING FILE:
       The number of observations in Using File equals the number of deaths in 
       the main CVSD file from 2006 to 2016.
  	
   - MERGE RESULT:
       Matched = These are death records from 2006 to 2016, with available 
         information on contributing causes in the Multiple Cause dataset.	
  	  
       Not matched from master = These are death records before 2006 and after 
         2016.
  	  
       Not matched from using = These are death records in the CVSD main file 
         with no information recorded on contributing causes in the Multiple 
         Cause file.
  * - - - - - - - - - - - - - - - - - - - - - - - - - - - - - - - - - - - - - */
			
  * Rename the 20 multi-cause variables to contributing cause
  rename ra_mc* icd_contr_cause*
	
  * Save as numeric if you can
  destring *, replace
	
  * Save
  save ".\Data\cvsd_2006to2016_MASTER.dta", replace
end	

* 8. PREPARE CENSUS 2006 (LONG-FORM)                                        ****
* ---------------------------------------------------------------------------- *
program define prep_cens
  * This program harmonizes the Census 2006 dataset. 
   
  * Open Census 2006 raw file
  use ".\Data\census_long_2006_RAW.dta", clear
		
  /* - - - - - - - - - - - - - - - - - - - - - - - - - - - - - - - - - - - - - *
     Note that the number of observations in this file is more than the number  
     of CanCHEC 2006 cohort, which equals the number of observations in Master 
     Keyfile. This is because not every respondent of the Census (long-form)  
     is eligible to be included in the CanCHEC cohort.
  * - - - - - - - - - - - - - - - - - - - - - - - - - - - - - - - - - - - - - */
	
  * Keep variables we need
  keep age hcdd cma sex hhinc_at nunits fsaname pcd pcsd pop pr prcdda       ///
    ruindfg rusize compw2 hhnum	ppnum nocsbrd dvismin lf71 sac_type
	
  * Rename variables and add a suffix of "cens" to those that we might have 
  * similars in other CanCHEC datasets
  rename age        age_cens
  rename cma        cma_cens
  rename sex        sex_cens
  rename hhinc_at   hhatinc
  rename nunits     hhsize
  rename fsaname    fsa_cens
  rename pcd        cd_cens
  rename pcsd       csd_cens
  rename pop        pop_csd
  rename pr         pr_cens
  rename ruindfg    rufg
  rename nocsbrd    occ
  rename compw2     w_cens			
			
  * Generate race variable based on CIHI methodology, using Visible Minority
  * variable available in Census.
  generate race = .
  replace race = 1 if inlist(dvismin,3)
  replace race = 2 if inlist(dvismin,1,9,10)
  replace race = 3 if inlist(dvismin,5)
  replace race = 4 if inlist(dvismin,7,8)
  replace race = 5 if inlist(dvismin,2)
  replace race = 6 if inlist(dvismin,4,6)
  replace race = 7 if inlist(dvismin,13)
  replace race = 8 if inlist(dvismin,15)
  replace race = 9 if inlist(dvismin,11)
  replace race = 10 if inlist(dvismin,12)

  *  Race and ethnicity in Census                                           **** 
  /* - - - - - - - - - - - - - - - - - - - - - - - - - - - - - - - - - - - - - *
  For dvismin == 14,  is  not included in the above categories.
    
  The following are the labels of values for the detailed visible minority 
  variable (dvismin) in Census:
  
    Visible Minority  |  Label
  --------------------+-----------------------------------------
                    1 |  Chinese
                    2 |  South Asian
                    3 |  Black
                    4 |  Filipino
                    5 |  Latin American
                    6 |  Southeast Asian
                    7 |  Arab
                    8 |  West Asian
                    9 |  Korean
                   10 |  Japanese
                   11 |  Visible minority, n.i.e.*
                   12 |  Multiple visible minority
                   13 |  Not a visible minority
                   14 |  Not applicable (Institutional resident)
                   15 |  Aboriginal self-reporting
  --------------------------------------------------------------
  Note: *'Visible minority, n.i.e.' includes respondents who reported a 
  write-in responses such as Guyanese, West Indian, Kurd, Tibetan, etc.
  
  The following are the labels of values for the race variable that is created 
  based on CIHI's methodology:		
  			
                 Race |  Label
  --------------------+---------------------------
                    1 |  Black
                    2 |  East Asian
                    3 |  Latin American
                    4 |  Middle Eastern
                    5 |  South Asian
                    6 |  Southeast Asian
                    7 |  White
                    8 |  Aboriginal
                    9 |  Other
                   10 |  Multiple visible minority
  ------------------------------------------------
  * - - - - - - - - - - - - - - - - - - - - - - - - - - - - - - - - - - - - - */                        
  
  * Generate education variable using the Highest Certificate, Diploma or 
  * Degree variable in Census
  generate edu = .
  replace edu = 1 if inlist(hcdd,1)
  replace edu = 2 if inlist(hcdd,2)
  replace edu = 3 if inlist(hcdd,3,4,5,6,7,8)
  replace edu = 4 if inlist(hcdd,9)
  replace edu = 5 if inlist(hcdd,10,11,12,13)
  replace edu = 6 if inlist(hcdd,14)
  
  * Generate employment variable using the Labour Force Activity - 1971 Census
  * Concepts variable
  generate emp = .
  replace emp = 1 if inlist(lf71,0)
  replace emp = 2 if inlist(lf71,1,2,3,4,5)
  replace emp = 3 if inlist(lf71,6,7)
  replace emp = 4 if inlist(lf71,8,9,10)
  
  * Generate household adjustment indicator to be used for adjusting after-tax 
  * household income by houshehold size	
  generate hhsize_adj = sqrt(hhsize)	
	
  * Generate income variable adjusted by household size
  generate hhatinc_adj = hhatinc / hhsize_adj		

  * Drop variables we don't need
  drop dvismin hcdd lf71 hhsize* hhatinc		

  * Save
  save ".\Data\census_long_2006_MASTER.dta", replace
	
end

* 9. CREATE NATIONAL COHORT                                                 ****
* ---------------------------------------------------------------------------- *
program define create_cohort_nat
  * This program links DAD (2006-16), CVSD(2006-16), and Census 2006, using 
  * CanCHEC key files, and produce one dataset which is event-based and covers 
  * all Canada.

  * The datasets that are linked by this program include DAD, CVSD, Census,
  * Master key file, Bootstrap key file, and CMDBonly key file.

  * Open harmonized Master key file
  use ".\Data\canchec2006_key_master_HARM.dta", clear
  
  * Merge with the Bootstrap key file to get the CanCHEC weight
  merge 1:1 uniqid using ".\Data\canchec2006_key_bs_HARM.dta", nogen 
  
  *  Merge results                                                          ****
  /* - - - - - - - - - - - - - - - - - - - - - - - - - - - - - - - - - - - - - *
   - MASTER FILE:
       The number of observations equals the number of people in the CanCHEC  
       2006 cohort.
     
   - USING FILE:
       The number of observations equals the number of people in the CanCHEC  
       2006 cohort.
  	
   - MERGE RESULT:
       Matched = All observations in using and master files should be matched.
  * - - - - - - - - - - - - - - - - - - - - - - - - - - - - - - - - - - - - - */
	
  * Merge with Census 2006
  merge 1:1 prcdda hhnum ppnum using ".\Data\census_long_2006_MASTER.dta",   ///
    nogen keep(3)
	
  *  Merge results                                                          ****
  /* - - - - - - - - - - - - - - - - - - - - - - - - - - - - - - - - - - - - - *
   - MASTER FILE:
       The number of observations equals the number of people in the CanCHEC  
       2006 cohort.
     
   - USING FILE:
       The number of observations equals the number of people in the Census 
	   2006.
  	
   - MERGE RESULT:
       Matched = The number of people in the CanCHEC 2006 cohort.	
	   
       Not matched from master = None.
	
       Not matched from using = The number of people in Census 2006 that did 
         not met the CanCHECS criteria to be included in the cohort. 
         E.g., institutional residents.
  * - - - - - - - - - - - - - - - - - - - - - - - - - - - - - - - - - - - - - */

  * Merge with CVSD (2006-16)
  merge m:1 event_year placeofdeath_province registration_number using       ///
    ".\Data\cvsd_2006to2016_MASTER.dta", nogen keep(1 3)

  *  Merge results                                                          ****
  /* - - - - - - - - - - - - - - - - - - - - - - - - - - - - - - - - - - - - - *
   - MASTER FILE:
       The number of observations equals the number of people in the CanCHEC  
       2006 cohort.
     
   - USING FILE:
       The number of observations equals the number of deaths recorded for 
       people in the CanCHEC 2006 cohort from 2006 to 2016.
  	
   - MERGE RESULT:
       Matched = The number of people in the CanCHEC 2006 cohort who died 
         between 2006 to 2016.	
	   
       Not matched from master = The number of people in CanCHEC 2006 who did 
         not die between 2006 to 2016.
	
       Not matched from using = The number of deaths happened to Canadians 
         between 2006 to 2016, who are not included in the CanCHEC 2006.
		 
    Note: Since there are no observations in our using file that have missing 
	  values for the three key variables, using m:1 did not make any 
          problem.		 
  * - - - - - - - - - - - - - - - - - - - - - - - - - - - - - - - - - - - - - */

  * Merge with CMDBonly file to add deaths that were not captured by CVSD
  merge 1:1 uniqid using ".\Data\canchec2006_key_cmdbonly_HARM.dta", nogen   ///
    update replace keep(1 4)

  *  Merge results                                                          ****
  /* - - - - - - - - - - - - - - - - - - - - - - - - - - - - - - - - - - - - - *
   - MASTER FILE:
       The number of observations equals the number of people in the CanCHEC 
       2006 cohort.
  
   - USING FILE:  
       The number of observations equals a subset of CMDB records for CanCHEC
       2006 deaths that do not link to a death record on the CVSD.
  	
   - MERGE RESULT:
  	Matched (missing updated) = The number of deaths that happened between
          2006 and 2016 among the CanCHEC 2006 cohort, which does not link to  
          a death record on the CVSD.
  * - - - - - - - - - - - - - - - - - - - - - - - - - - - - - - - - - - - - - */	
		
  * Merge with DAD (2006-16)
  merge 1:m uniqid using ".\Data\canchec2006_dad_2006to2016_MASTER.dta", nogen

  *  Merge results                                                          ****
  /* - - - - - - - - - - - - - - - - - - - - - - - - - - - - - - - - - - - - - *
   - MASTER FILE:
       The number of observations equals the number of people in the CanCHEC 
       2006 cohort.

   - USING FILE:  
       The number of observations equals the number of hospitalizations 
         recorded for people in the CanCHEC 2006 cohort between 2006 and 2016.

   - MERGE RESULT:
       Not matched from master = The number of people in CanCHEC 2006 cohort  
         who have no hospitalization records in DAD between 2006 and 2016.
  	
       Not matched from using = Zero.
  	
  NOTE: After this merge, the person-based dataset is changed to an event-based
  	dataset. This event-based dataset has 9,625,282 observations.
  * - - - - - - - - - - - - - - - - - - - - - - - - - - - - - - - - - - - - - - */
	
  * Rename variables
  rename prcdda            da_06
  rename event_year        year_death
  rename icd_contr_cause*  icd_contr_cause_* 

  * Save
  save ".\Data\canchec2006_cohort_can_06to16_e_USE.dta", replace
		
end
	
* 10. CREATE PROVINCIAL COHORT (SASKATCHEWAN)                               ****
* ---------------------------------------------------------------------------- *
program define create_cohort_pr
  * This program creates a cohort of Saskatchewanians by using the Cohort of 
  * Canadians (CanCHEC2006_cohort_can_06to16_e_USE) created in the previous 
  * step.

  * We set the following criteria to identify Saskatchewanians in the national 
  * cohort:
  *   1. In DAD: having an SK health card OR postal code starting with S
  *   2. In CVSD: province of residence is SK OR postal code starts with S
  *   3. In Census: province of residence is SK OR postal code starts with S
  * If an individual in our national cohort meets AT LEAST one criterion, we 
  * identified them as a Saskatchewanian and included them in the provincial 
  * Cohort.

  * THIS SECTION OF THE CODE CAN BE CHANGED TO GENERATE COHORTS FOR OTHER 
  * PROVINCES AND TERRITORIES.   

  * Open event-based Canadian cohort
  use ".\Data\canchec2006_cohort_can_06to16_e_USE.dta", clear
  
  * Generate a dummy variable that identifies Saskatchewanian abstracts
  generate resdn_sk_dad_e = 0
	
  * Identify Saskatchewanian abstracts (criteria: SK health card OR postal 
  * code starts with S)
  replace resdn_sk_dad_e = 1 if hcard_pr == "SK" |                          ///
                                regexm(resdn_pcode_dad,"^S")	
	
  * Identify Saskatchewanians based on having at least one Saskatchewanian 
  * abstract
  egen resdn_sk_dad_p = max(resdn_sk_dad_e), by(uniqid)	
  
  * Drop the event-level variable that we no longer need
  drop resdn_sk_dad_e
		
  * Generate a dummy variable that identifies Saskatchewanian deaths
  generate resdn_sk_cvsd_p = 0
	
  * Identify Saskatchewanian deaths (criteria: province of residence OR postal 
  * code starts with S)	
  replace resdn_sk_cvsd_p = 1 if resdn_pr_cvsd == 947 |                      ///
                                 regexm(resdn_pcode_cvsd,"^S")
		
  * Generate a dummy variable that identifies Saskatchewanians in census
  generate resdn_sk_cens_p = 0
	
  * Identify Saskatchewanians in census (criteria: province of residence OR 
  * postal code starts with S)	
  replace resdn_sk_cens_p = 1 if pr_cens == 47 |                             /// 
                                 regexm(fsa_cens,"^S")

  * Identify Saskatchewanians according to DAD, CVSD, and Census information
  generate resdn_sk_p = 0 
  replace resdn_sk_p = 1 if resdn_sk_cvsd_p == 1 |                           ///
                            resdn_sk_dad_p == 1 |                            ///
                            resdn_sk_cens_p == 1
	
  * Drop non-Saskatchewanians and create a cohort of Saskatchewan
  keep if resdn_sk_p == 1		
		
  * Save
  save ".\Data\canchec2006_cohort_sk_06to16_e_USE.dta", replace	

end 

* 11. IDENTIFY SUBSTANCE USE HARM EVENTS AND PESUH                          ****
* ---------------------------------------------------------------------------- *
program define find_hosp_death_all  
  * This program identifies hospitalization and death events overall (due to 
  * any reason.)

  * Details on how variables are named in this program                      ****
  /* - - - - - - - - - - - - - - - - - - - - - - - - - - - - - - - - - - - - - *
	The prefix "hosp" indicates a dummy variable for hospitalization.
	The prefix "death" indicates a dummy variable for a death event.
    The suffix "_e" indicates an event-level variable.
    The suffix "_p" indicates a person-level variable.    
  * - - - - - - - - - - - - - - - - - - - - - - - - - - - - - - - - - - - - - */
		
  * Open the cohort of Saskatchewan
  use ".\Data\canchec2006_cohort_sk_06to16_e_USE.dta", clear
	
  * Create a dummy variable for hospitalization events
  generate hosp_e = 0
  replace hosp_e = 1 if !missing(dad_transaction_id)
	
  * Create a dummy variable for individuals who were hospitalized at least once
  * between 2006-16
  egen hosp_p = max(hosp_e), by(uniqid)
	
  * Create a dummy variable for death events
  generate death_e = 0
  replace death_e = 1 if !missing(icd_underly_cause)
	
  * Create a dummy to identify individuals who died between 2006 to 2016	
  egen death_p = max(death_e), by(uniqid)

  /* - - - - - - - - - - - - - - - - - - - - - - - - - - - - - - - - - - - - - *
  NOTE: While the sum of hosp_e variable will get the number of 
        hospitalizations, this is not true for death. To get the number of 
        deaths, the dataset should be collapsed to the individual level.
  * - - - - - - - - - - - - - - - - - - - - - - - - - - - - - - - - - - - - - */
  
  * Save
  save ".\Data\canchec2006_cohort_sk_06to16_all_hospdeath_e_MASTER.dta", replace
end

program define find_hosp_su
  * This program identifies hospitalization events due to substance use (SU)  
  * and people who experienced hospitalization due to substance use between 
  * 2006 to 2016 using an algorithm developed by CIHI.

  * About CIHI's methodology in finding hospitalization due to SU           ****
  /* - - - - - - - - - - - - - - - - - - - - - - - - - - - - - - - - - - - - - *
     In DAD, the diagnosis information of a single hospitalization event is 
     recorded in 4 sets of variables: Diagnosis Prefix, Diagnosis Code, 
     Diagnosis Type, and Diagnosis Cluster. There must be at least one 
     diagnosis per abstract. There could be up to 25 diagnoses recorded per 
     abstract.

     CIHI categorizes hospitalizations due to substance use by type of 
     hospitalization (Mental and behavioural disorders, Poisoning, and Medical
     condition and external cause) and by substance category (Alcohol, Opioids,
     Cannabis, Other CNS depressants, Cocaine, Other CNS stimulants, Unknown 
     and multiple substances, and Other substances.)
     
     CIHI uses the Diagnosis Code and Diagnosis Type variables (25 variables
     per each) to find hospitalizations due to substance use. Following is the
     description of these two sets of variables:
	 
       1. Diagnosis Code: This variable is the ICD-10-CA classification code 
          that describes the diagnoses/conditions of the patient during the 
          length of stay in the health care facility. There are at least one 
          and up to 25 Diagnosis Codes recorded for each hospitalization 
          (diag_code_`i' for 1 to 25). 
	   
       2. Diagnosis Type: This variable is an alpha or a numeric code meant to 
          signify the impact that the condition had on the patient’s care as 
          evidenced in the physician/allied health care provider documentation.
	
               Diagnosis Type |  Title
          --------------------+------------------------------------------
                            M |  Most Responsible Diagnosis (MRDx)
                            1 |  Pre-admit Comorbidity
                            2 |  Post-admit Comorbidity
                            3 |  Secondary Diagnosis
                      W, X, Y |  Service Transfer Diagnosis	
                            4 |  Morphology Code
                            5 |  Admitting Diagnosis
                            6 |  Proxy Most Responsible Diagnosis (MRDx)
                         7, 8 |  Restricted to CIHI
                            9 |  External Cause of Injury Code
                            0 |  Newborn
          ---------------------------------------------------------------
	  
     CIHI considers a hospitalization related to substance use if a 
     hospitalization abstract has a diagnosis code listed in the table 
     provided in Appendix B AND the diagnosis type associated with that 
     diagnosis code is "M","1","2","W","X","Y",or "9". We call this condition 
     GENERAL CRITERIA.
    
     There are 3 sets of ICD codes that should meet EXTRA CONDITIONS, as 
     follows:
       1. O993 should be included only if the corresponding mental and 
          behavioural disorders ICDs as diagnosis type (3) is in the same 
          abstract. That means, F10 for Alcohol, F11 for Opioids, F12 for 
          Cannabis, F13 for Other CNS depressants, F14 for Cocaine, F15 for 
          Other CNS stimulants, F19 for unknown and multiple substances, and 
          F16 and F18 for Other Substances.
       2. For Unkown and multiple substances category, X41, X61, and Y11 
          should be included if neither T42.– nor T43.– are in the same 
          abstract.
       3. For Unkown and multiple substances category, X42, X62, and Y12 
          should be included if T40.– is not in the same abstract.   
  * - - - - - - - - - - - - - - - - - - - - - - - - - - - - - - - - - - - - - */
  
  * Details on how variables are named in this program                      ****
  /* - - - - - - - - - - - - - - - - - - - - - - - - - - - - - - - - - - - - - *
     The prefix "hosp" indicates a dummy variable for hospitalization.
     The prefix "death" indicates a dummy variable for a death event.
     The suffix "_e" indicates an event-level variable.
     The suffix "_p" indicates a person-level variable.    
  * - - - - - - - - - - - - - - - - - - - - - - - - - - - - - - - - - - - - - */
  
  * Open event-based provincial dataset with identified OVERALL 
  * hospitalizations and death	
  use ".\Data\canchec2006_cohort_sk_06to16_all_hospdeath_e_MASTER.dta", clear	
  
  * Save ICD-10 codes for hospitalizations related to Alcohol for those 
  * conditions that should meet the GENERAL CRITERIA in a local macro 		
  local hosp_alc_gen                                                         ///
        F10 T51 E244 G312 G621 G721 I426 K292 K70 K852 K860 O354 Q860 R780   ///
        X45 X65 Y15

  * Save ICD-10 codes for hospitalizations related to Alcohol for those 
  * conditions that should meet the EXTRA CRITERIA in a local macro 			
  local hosp_alc_o993                                                        ///
        F10

  * Save ICD-10 codes for hospitalizations related to Opioids for those 
  * conditions that should meet the GENERAL CRITERIA in a local macro 		
  local hosp_opi_gen                                                         ///
        F11 T400 T401 T402 T403 T404 T406 			

  * Save ICD-10 codes for hospitalizations related to Opioids for those 
  * conditions that should meet the EXTRA CRITERIA in a local macro 
  local hosp_opi_o993                                                        ///
        F11
 																						
  * Save ICD-10 codes for hospitalizations related to Cannabis for those 
  * conditions that should meet the GENERAL CRITERIA in a local macro 		
  local hosp_can_gen                                                         ///
        F12 T407			

  * Save ICD-10 codes for hospitalizations related to Cannabis for those 
  * conditions that should meet the EXTRA CRITERIA in a local macro   
  local hosp_can_o993                                                        ///
        F12
																					
  * Save ICD-10 codes for hospitalizations related to Other CNS depressants for  
  * those conditions that should meet the GENERAL CRITERIA in a local macro 		
  local hosp_cnsdep_gen                                                      ///
        F13 T423 T424 T426 T427			

  * Save ICD-10 codes for hospitalizations related to Other CNS depressants for  
  * those conditions that should meet the EXTRA CRITERIA in a local macro   
  local hosp_cnsdep_o993                                                     ///
        F13
		
  * Save ICD-10 codes for hospitalizations related to Cocaine for those 
  * conditions that should meet the GENERAL CRITERIA in a local macro 		
  local hosp_coc_gen                                                         ///
        F14 T405			

  * Save ICD-10 codes for hospitalizations related to Cocaine for those 
  * conditions that should meet the EXTRA CRITERIA in a local macro   
  local hosp_coc_o993                                                        ///
        F14	
		
  * Save ICD-10 codes for hospitalizations related to Other CNS stimulants for  
  * those conditions that should meet the GENERAL CRITERIA in a local macro 		
  local hosp_cnsstim_gen                                                     ///
        F15 T436			

  * Save ICD-10 codes for hospitalizations related to Other CNS stimulants for  
  * those conditions that should meet the EXTRA CRITERIA in a local macro   
  local hosp_cnsstim_o993                                                    ///
        F15
		
  * Save ICD-10 codes for hospitalizations related to Unkown and multiple 
  * substances for those conditions that should meet the GENERAL CRITERIA in a 
  * local macro 		
  local hosp_unkmult_gen                                                     ///
        F19 T438 T439 O355			

  * Save ICD-10 codes for hospitalizations related to Unkown and multiple 
  * substances for those conditions that should meet the EXTRA CRITERIA in a 
  * local macro   
  local hosp_unkmult_o993                                                    ///
        F19
		
  * Save ICD-10 codes for hospitalizations related to Other substances for  
  * those conditions that should meet the GENERAL CRITERIA in a local macro 		
  local hosp_other_gen                                                       ///
        F16 F18 F55 T408 T409			

  * Save ICD-10 codes for hospitalizations related to Other substances for  
  * those conditions that should meet the EXTRA CRITERIA in a local macro   
  local hosp_other_o993                                                      ///
        F16 F18	
 
  * Start a loop to identify SU hospitalizations for those with only GENERAL
  * CRITERIA and those with O993 EXTRA CRITERIA, by substance category
  foreach category in alc opi can cnsdep coc cnsstim unkmult other {			
    
    * Generate dummy variable for hospitalizations, by substance category
    gen hosp_`category'_e = 0

    * Check the hospitalization records against our SU case-fiding algorithm
    * (Appendix B) using 25 Diagnosis Code and 25 Diagnosis type variables
    forvalues i = 1/25 {
      
      * Start a loop to find SU hospitalizations with GENERAL CRITERIA
      foreach icd of local hosp_`category'_gen{
    
        * Replace SU hospitalization dummy with 1 if the abstract meets the
        * GENERAL CRITERIA 
        replace hosp_`category'_e = 1                                        ///
          if regexm(diag_code_`i',"^`icd'") &                                ///
             inlist(diag_type_`i',"M","1","2","W","X","Y","9")
      }  
    }	
	
    * Generate dummy variable for hospitalization due to O993 for all categories
    gen hosp_`category'_o993 = 0
          
    * Start a loop to find SU hospitalizations due to O993 and save it for all 
    * categories
    forvalues i = 1/25 {
      replace hosp_`category'_o993 = 1                                       ///
        if regexm(diag_code_`i',"^O993") &                                   ///
           inlist(diag_type_`i',"M","1","2","W","X","Y","9")  
    }
    
    * Replace SU hospitalization dummy with 1 if the abstract with O993 meets
    * the EXTRA CONDITIONS
    forvalues i = 1/25 {
      foreach icd of local hosp_`category'_o993 {
        replace hosp_`category'_e = 1                                        ///
          if hosp_`category'_o993 == 1 &                                     ///
    	     regexm(diag_code_`i',"^`icd'") &                                ///
             diag_type_`i' == "3"
      }   
    }
  }

  * Generate dummy variables for hospitalization due to conditions related to
  * Uknown and Multiple Substances category, that should meet EXTRA CRITERIA
  gen hosp_unkmult_x41 = 0
  gen hosp_unkmult_x42 = 0 
 
  * Start a loop to find SU hospitalizations with EXTRA CRITERIA (except O993)
  forvalues i = 1/25 {
    
    * Find hospitalizations due to X41, X61, and Y11 
    foreach icd in X41 X61 Y11 {
      replace hosp_unkmult_x41 = 1                                           /// 
        if regexm(diag_code_`i',"^`icd'") &                                  ///
           inlist(diag_type_`i',"M","1","2","W","X","Y","9")  
    }
	
    * Find hospitalizations due to X62 X42 Y12 
    foreach icd in X62 X42 Y12 {
      replace hosp_unkmult_x42 = 1                                           ///
        if regexm(diag_code_`i',"^`icd'") &                                  ///
           inlist(diag_type_`i',"M","1","2","W","X","Y","9")  
    }
  }	
 
  * Replace unkmult hospitalization dummies with 0 if the abstracts related to 
  * X41, X61, Y11, X62, X42, and Y12 does not meet the EXTRA CRITERIA
  forvalues i = 1/25 {
    
    * For X41, X61, and Y11
    replace hosp_unkmult_x41 = 0 if regexm(diag_code_`i',"^T42") |           ///
                                    regexm(diag_code_`i',"^T43")

    * For X62, X42, and Y12									
    replace hosp_unkmult_x42 = 0 if regexm(diag_code_`i',"^T40")   
  } 

   * Replace SU hospitalization dummy with 1, if the Unkmult dummies for those
   * with EXTRA CRITERIA are 1
   replace hosp_unkmult_e = 1 if hosp_unkmult_x41 == 1 |                    ///
                                 hosp_unkmult_x42 == 1
	
   * Generate a single dummy variable for all SU hospitalizations
   generate hosp_su_e = 0
	
   * Replace SU hospitalization dummy with one if any of the eight SU 
   * categories were identified as an SU case for an abstract
   replace hosp_su_e = 1 if hosp_alc_e == 1 |                                ///
                            hosp_opi_e == 1 |                                ///
                            hosp_can_e == 1 |                                ///
                            hosp_cnsdep_e == 1 |                             ///
                            hosp_coc_e == 1 |                                ///
                            hosp_cnsstim_e == 1 |                            ///
                            hosp_unkmult_e == 1 |                            ///
                            hosp_other_e == 1                               
			
  * Generate dummy variables for individuals who had experienced at least 
  * one SU hospitalization between 2006 and 2016, by SU category
  foreach category in alc opi can cnsdep coc cnsstim unkmult other {
    egen hosp_`category'_p = max(hosp_`category'_e), by(uniqid)
  }
  
  * Generate a dummy variable for individuals who had experienced at least
  * one SU hospitalization between 2006 and 2016
  egen hosp_su_p = max(hosp_su_e), by(uniqid)
  
  * Drop variables we don't need
  drop *_o993 *_x41 *_x42
  	
  * Save
  save                                                                       ///
    ".\Data\canchec2006_cohort_sk_06to16_all_hospdeath_su_hosp_e_MASTER.dta" ///
    , replace	
end 

program define find_death_su	
  * This program identifies death events due to substance use using an  
  * algorithm developed for this study. The algorithm is provided in  
  * Appendix B, which is based on related literature.

  *  About CIHI's methodology in finding hospitalization due to SU          ****
  /* - - - - - - - - - - - - - - - - - - - - - - - - - - - - - - - - - - - - - *
     To identify deaths due to SU for non-poisoning cases, we simply looked at 
     the underlying cause of death to see if it matches the ICD code
     listed in our SU case-finding algorithm (Appendix B.) The underlying 
     cause of death is a variable in the main CVSD datasets named 
     "death_cause_4digits". This variable contains the ICD code attributed to 
     the cause of death.
	 
     For poisoning cases of deaths due to SU, however, we took a different 
     approach. To identify these deaths, we examined the CVSD Multiple Cause
     datasets in addition to the main files. The contributing cause information 
     is included in a set of 20 variables named "ra_mc_`i'", where i is 1 to 20. 
     These variables provide additional information for each death registered 
     in the main CVSD files, which are needed to identify SU poisoning cases.
     The case-finding algorithm for these types of deaths is provided in 
     Appendix B.
	 
     For poisoning cases, we need the contributing cause of death to identify
     the substance causing the poisoning. The underlying cause only indicates
     if a death was due to poisoning. By relying on contributing causes of 
     death, we can determine if the poisoning was due to a substance of our
     interest and categorize it into our eight substance categories.
  * - - - - - - - - - - - - - - - - - - - - - - - - - - - - - - - - - - - - - */
  
  * Open event-based provincial dataset with identified OVERALL hospitalizations
  * and deat and SU hospitalizations
  use                                                                        ///
    ".\Data\canchec2006_cohort_sk_06to16_all_hospdeath_su_hosp_e_MASTER.dta" ///
    , clear	
  
  * Save ICD-10 codes for underlying cause of SU deaths (not poisoning) by 
  * substance category in local macros
  local alc_notpois                                                          ///
        F10 E244 G312 G621 G721 I426 K292 K70 K852 K860	O354 Q860 R780 X45   ///
        X65 Y15
		
  local opi_notpois                                                          ///
        F11 
		
  local can_notpois                                                          ///
        F12
		
  local cnsdep_notpois                                                       ///
        F13
		
  local coc_notpois                                                          ///
        F14	
		
  local cnsstim_notpois                                                      ///
        F15
		
  local unkmult_notpois                                                      ///
        F19 X41 X42 X61 X62 Y11 Y12
		
  local other_notpois                                                        ///
        F16 F18 F55 		
  
  * Save ICD-10 codes for the underlying cause of SU death (poisoning) in a  
  * local macro (all categories except Opioids)
  local pois_under                                                           ///
        X40 X41 X42 X43 X44 X60 X61 X62 X63 X64 X85 Y10 Y11 Y12 Y13 Y14

  * Save ICD-10 codes for the underlying cause of SU death (poisoning) in a  
  * local macro (Opioids)
  local pois_under_opi                                                       ///
        X40 X41 X42 X43 X44 X60 X61 X62 X63 X64 X85 Y11 Y12 Y13 Y14

  * Save ICD-10 codes for contributing causes of death related to substance 
  * use (poisoning) by category  
  local alc_pois_contr                                                       ///
        T51
		
  local opi_pois_contr                                                       ///
        T400 T401 T402 T403 T404 T406
  
  local can_pois_contr                                                       ///
        T407
		
  local cnsdep_pois_contr                                                    ///
        T423 T424 T426 T427
		
  local coc_pois_contr                                                       ///
        T405
		
  local cnsstim_pois_contr                                                   ///
        T436
		
  local unkmult_pois_contr                                                   ///
        T438 T439
		
  local other_pois_contr                                                     ///
        T408 T409
			
  * Identify SU deaths (non-poisoning) using the underlying cause of death
  foreach category in alc opi can cnsdep coc cnsstim unkmult other {

    * Generate a dummy variable for non-poisonous death by substance category
    generate death_`category'_notpois_e = 0
    
    * Use ICD codes saved in local macros to identify SU deaths
    foreach icd of local `category'_notpois {
      replace death_`category'_notpois_e = 1 if regexm(icd_underly_cause, "^`icd'")
    }
  }

  * Identify deaths that meet the underlying cause criteria for all categories 
  * (except for Opioids) poisoning death
  foreach category in alc can cnsdep coc cnsstim unkmult other {

    * Generate a dummy variable for the underlying cause criteria of poisonous 
    * deaths
    generate `category'_pois_under = 0

    * Use ICD codes saved in local macros to find abstracts meeting the 
    * underlying cause criteria for poisonous death
    foreach icd of local pois_under {
      replace `category'_pois_under = 1 if regexm(icd_underly_cause,"^`icd'")
    }
  }

  * Identify deaths that meet the underlying cause criteria for Opioids 
  * poisoning death by creating a dummy variable
  generate opi_pois_under = 0

  * Use ICD codes saved in a local macro to find abstracts meeting the criteria
  foreach icd of local pois_under_opi {
    replace opi_pois_under = 1 if regexm(icd_underly_cause,"^`icd'")
  }

  * Identify SU deaths (poisoning) using both underlying and contributing 
  * cause of death information
  foreach category in alc opi can cnsdep coc cnsstim unkmult other {
    
    * Generate a dummy variable for poisonous death by category
    generate death_`category'_pois_e = 0

    * Start a loop to grab information from the 20 variables containing 
    * information on contributing causes of death
    forvalues n = 1/20 {

      * Use ICD codes saved in local macros for contributing causes of 
      * death criteria      
      foreach icd of local pois_contr_`category'{
        * Use both underlying and contributing causes of death information 
        * to find SU poisonous deaths
        replace death_`category'_pois_e = 1                               ///
          if `category'_pois_under == 1 &                                 ///
             regexm(icd_contr_cause_`n', "^`icd'") 
      }       
    }
  } 

  * Generate a dummy variable to indicate if a death is identified as a SU
  * death or not (poisoning or not poisoning) by substance category
  foreach category in alc opi can cnsdep coc cnsstim unkmult other {
    generate death_`category'_e = 0
    replace death_`category'_e = 1 if death_`category'_pois_e == 1 |     ///
                                      death_`category'_notpois_e == 1
  } 

  * Create a single dummy variable for all SU deaths
  generate death_su_e = 0
	
  * Replace SU death dummy with one if any of the eight substance 
  * categories were identified as an SU case for a death
  replace death_su_e = 1 if death_alc_e == 1 |                          ///
                            death_opi_e == 1 |                          ///
                            death_can_e == 1 |                          ///
                            death_cnsdep_e == 1 |                       ///
                            death_coc_e == 1 |                          ///
                            death_cnsstim_e == 1 |                      ///
                            death_unkmult_e == 1 |                      ///
                            death_other_e == 1 

  * Generate dummy variables for individuals who died due to SU between 
  * 2006 and 2016, by substance category
  foreach category in alc opi can cnsdep coc cnsstim unkmult other {
    egen death_`category'_p = max(death_`category'_e), by(uniqid)
  }
  
  * Generate a dummy variable for individuals who died du to SU between
  * 2006 and 2016
  egen death_su_p = max(death_su_e), by(uniqid)                          
					
  * Create a dummy variable to identify people who experienced at least 
  * one substance use harm event (hospitalization or death) during 
  * 2006-16 (PESUH)
  generate pesuh = 0
  replace pesuh = 1 if hosp_su_p == 1 |                                 ///
                       death_su_p == 1 
			
  * Drop variable we don't need
	drop *pois_e *_under

  * Save
  save                                                                        ///
    ".\Data\canchec2006_cohort_sk_06to16_all_hospdeath_su_hospdeath_e_USE.dta", /// 
	replace			
end
	
program define label_vars
  * Since the datasets used in this do-file have large size, where possible,  
  * we change the string variables to integers to decrease the file sizes, and 
  * therefore, speed up the execution time. We label all variables at the end 
  * to facilitate working with data and understanding it.

  * Open non-labeled substance use cohort dataset
  use                                                                        ///
    ".\Data\canchec2006_cohort_sk_06to16_all_hospdeath_su_hospdeath_e_USE.dta" ///
	, clear
	
  * Label variables
  label variable  uniqid                  "Unique ID (CanCHEC 2006)"
  label variable  da_06                   "dissemination area (prcdda in CanCHEC 2006)"
  label variable  hhnum                   "key for household table (CanCHEC 2006)"
  label variable  ppnum                   "key for person table (CanCHEC 2006)"
  label variable  pp_id                   "alternative key for census person table (CanCHEC 2006)"
  label variable  year_death              "year when death occurred (event year in CanCHEC 2006)"
  label variable  placeofdeath_province   "province where death occurred (CanCHEC 2006)"
  label variable  registration_number     "death registration number (CanCHEC 2006)"
  label variable  w_cohort                "CanCHEC 2006 weight (CanCHEC 2006)"
  label variable  age_cens                "age (census 2006)"
  label variable  cma_cens                "census metropolitan area or census agglomeration of current residence (2006)"
  label variable  w_cens                  "composite weight (perswt + occwtp -1) (census 2006)"
  label variable  fsa_cens                "forward sortation area (census 2006)"
  label variable  occ                     "labour market activities : occupation broad categories (census 2006)"
  label variable  cd_cens                 "census division of current residence (census 2006)"
  label variable  csd_cens                "census subdivision of current residence (census 2006)"
  label variable  pop_csd                 "population size group of current census subdivision of residence (census 2006)"
  label variable  pr_cens                 "province or territory of current residence (census 2006)"
  label variable  rufg                    "rural, urban classification (census 2006)"
  label variable  rusize                  "rural, urban size code (census 2006)"
  label variable  sac_type                "statistical area classification (census 2006)"
  label variable  sex_cens                "sex (census 2006)"
  label variable  race                    "ethnicity (census 2006)"
  label variable  edu                     "education (census 2006)"
  label variable  emp                     "employment status (census 2006)"
  label variable  hhatinc_adj             "household after-tax income adjusted for household size (census 2006)"
  label variable  age_death               "age of deceased (CVSD)"
  label variable  resdn_pr_cvsd           "usual residence of deceased: province (CVSD)"
  label variable  resdn_pcode_cvsd        "usual residence of deceased: postal code (CVSD)"
  label variable  icd_underly_cause       "ICD for underlying cause of death (CVSD)"
  label variable  sex_death               "sex of deceased (CVSD)"
  label variable  dad_transaction_id      "DAD transaction ID (CanCHEC 2006)"
  label variable  submitting_prov_code    "submitting province code (DAD)"
  label variable  inst_code               "institution number (DAD)"
  label variable  year_hosp               "fiscal year of hospitalization (DAD)"
  label variable  hcard_pr                "province issuing the health card (DAD)"
  label variable  resdn_pcode_dad         "patient postal code (DAD)"
  label variable  age_hosp                "age at the time of hospitalization (DAD)"
  label variable  admission_date          "admission date (DAD)"
  label variable  discharge_date          "discharge date (DAD)"
  label variable  total_los_days          "total length of stay (DAD)"
  label variable  acute_los_days          "acute length of stay (DAD)"
  label variable  alc_los_days            "alternative level of care length of stay (DAD)"
  label variable  sex_hosp                "sex of patient (DAD)"
  label variable  resdn_sk_dad_p          "resident of Saskatchewan based on DAD"
  label variable  resdn_sk_cvsd_p         "resident of Saskatchewan based on CVSD"
  label variable  resdn_sk_cens_p         "resident of Saskatchewan based on census 2006"
  label variable  resdn_sk_p              "person residing in SK at some point between 2006 and 2016"
  label variable  hosp_e                  "hospitalization event"
  label variable  hosp_p                  "hospitalized person"
  label variable  death_e                 "death event"
  label variable  death_p                 "deceased person"
  label variable  hosp_alc_e              "hospitalization event due to alcohol"
  label variable  hosp_opi_e              "hospitalization event due to opioid"
  label variable  hosp_can_e              "hospitalization event due to cannabis"
  label variable  hosp_cnsdep_e           "hospitalization event due to other CNS depressants"
  label variable  hosp_coc_e              "hospitalization event due to cocaine"
  label variable  hosp_cnsstim_e          "hospitalization event due to other CNS stimulants"
  label variable  hosp_unkmult_e          "hospitalization event due to unknown and multiple substances"
  label variable  hosp_other_e            "hospitalization event due to other substances"
  label variable  hosp_su_e               "hospitalization event due to substance use"
  label variable  hosp_alc_p              "person hospitalized due to alcohol"
  label variable  hosp_opi_p              "person hospitalized due to opioids"
  label variable  hosp_can_p              "person hospitalized due to cannabis"
  label variable  hosp_cnsdep_p           "person hospitalized due to other CNS depressants"
  label variable  hosp_coc_p              "person hospitalized due to cocaine"
  label variable  hosp_cnsstim_p          "person hospitalized due to other CNS stimulants"
  label variable  hosp_unkmult_p          "person hospitalized due to unknown and multiple substances"
  label variable  hosp_other_p            "person hospitalized due to other substances"
  label variable  hosp_su_p               "person hospitalized due to substance use"
  label variable  death_alc_e             "death event due to alcohol"
  label variable  death_opi_e             "death event due to opioids"
  label variable  death_can_e             "death event due to cannabis"
  label variable  death_cnsdep_e          "death event due to other CNS depressants"
  label variable  death_coc_e             "death event due to cocaine"
  label variable  death_cnsstim_e         "death event due to other CNS stimulants"
  label variable  death_unkmult_e         "death event due to unknown and multiple substances"
  label variable  death_other_e           "death event due to other substances"
  label variable  death_su_e              "death event due to substance use"
  label variable  death_alc_p             "person died due to alcohol"
  label variable  death_opi_p             "person died due to opioids"
  label variable  death_can_p             "person died due to cannabis"
  label variable  death_cnsdep_p          "person died due to other CNS depressants"
  label variable  death_coc_p             "person died due to cocaine"
  label variable  death_cnsstim_p         "person died due to other CNS stimulants"
  label variable  death_unkmult_p         "person died due to unknown and multiple substances"
  label variable  death_other_p           "person died due to other substances"
  label variable  death_su_p              "person died due to substance use"
  label variable  pesuh                   "person who experienced substance use harm"
  
  * Label contributing causes of death variables
  forvalues i = 1/20 {
    label variable icd_contr_cause_`i' "ICD for contributing cause of death `i' (CVSD)"
  }
  
  * Label diagnosis cluster, prefix, code, and type variables
  forvalues i = 1/25 {
    label variable diag_cluster_`i' "diagnosis cluster `i' (DAD)"
    label variable diag_prefix_`i'  "diagnosis prefix `i' (DAD)"
    label variable diag_code_`i'    "diagnosis code `i' (DAD)"
    label variable diag_type_`i'    "diagnosis type `i' (DAD)"	
  }
  
  * Label census sex variables' values
  label define sexfull                                                       ///
    1  "Female"                                                              ///
    2  "Male", replace          
  label values sex_cens sexfull
  
  * Label DAD sex variable's values
  label define sexfull                                                       ///
    0  "Other"                                                               ///
    1  "Female"                                                              ///
    2  "Male", replace  
  label values sex_hosp sexfull	
  
  * Label CVSD sex variable's values
  label define sexfull                                                       ///
    0  "Unknown"                                                             ///
    1  "Female"                                                              ///
    2  "Male", replace  
  label values sex_death sexfull	
  
  * Label the generated race variable
  label define racefull                                                      ///
    1 "Black"                                                                ///
    2 "East Asian"                                                           ///
    3 "Latin American"                                                       ///
    4 "Middle Eastern"                                                       ///
    5 "South Asian"                                                          ///
    6 "Southeast Asian"                                                      ///
    7 "White"                                                                ///
    8 "Aboriginal"                                                           ///
    9 "Other"                                                                ///
    10 "Multiple visible minority", replace          
  label values race racefull       
    
 * Label the generated education variable
  label define education                                                     ///
    1  "Less than high school"                                               ///
    2  "High school graduation certificate or equivalency certificate"       ///
    3  "Non-university post-secondary certificate or diploma"                ///
    4  "Bachelor’s degree"                                                   ///
    5  "University certificate or diploma above bachelor level"              ///
    6  "Not applicable (Institutional residents)", replace               
  label values edu education
  	
  * Label the generated employment variable
  label define employment                                                    ///
    1  "Not applicable, less than 15 years"                                  ///
    2  "Employed"                                                            ///
    3  "Not in labour force"                                                 ///
    4  "Unemployed", replace                                                     
  label values emp employment 	
  
  * Label all derived dummy variables 
  label define dummy                                                         ///
    0  "No"                                                                  ///
    1  "Yes", replace
  label values hosp_* dummy
  label values death_* dummy
  label values resdn_sk_* dummy
  label values pesuh dummy
  
  * Save labeled substance use cohort dataset
  save                                                                       ///
    ".\Data\canchec2006_cohort_sk_06to16_all_hospdeath_su_hospdeath_e_FINAL.dta" ///
	, replace
end	

* A1. CONTROL
* ---------------------------------------------------------------------------- *
* Rename raw data files
  move_raw
  
* Harmonize CanCHEC 2006 keyfiles
  ** Harmonize Master keyfile
  harm_keyfile_master
  
  ** Harmonize bootstrap keyfile
  harm_keyfile_bs
  
  ** Harmonize CMDBonly keyfile
  harm_keyfile_cmdbonly
  
* Prepare DAD
  ** Reduce DAD annual files
  reduce_dad
  
  ** Append DAD annual files and link to DAD keyfile
  append_dad

* Prepare CVSD
  ** Reduce CVSD files
  reduce_cvsd

  ** Append CVSD annual files
  append_cvsd

  ** Merge CVSD main file to multi cause file
  merge_cvsd_multi

* Prepare Census
  prep_cens  

* Create national cohort
  create_cohort_nat

* Create provincial cohort
  create_cohort_pr

* Identify SUH and PESUH
  ** Find hospitalizations and deaths due to any reason
  find_hosp_death_all

  ** Find hospitalizations due to SU
  find_hosp_su 

  ** Find deaths due to SU
  find_death_su

  ** Label variables and values
  label_vars
```
